# Patients with fibrosis from non-alcoholic steatohepatitis have heterogeneous intrahepatic macrophages and therapeutic targets

**DOI:** 10.1101/2023.02.16.23285924

**Authors:** Omar A. Saldarriaga, Santhoshi Krishnan, Timothy G. Wanninger, Morgan Oneka, Arvind Rao, Daniel Bao, Esteban Arroyave, Joseph Gosnell, Michael Kueht, Akshata Moghe, Daniel Millian, Jingjing Jiao, Jessica I. Sanchez, Heidi Spratt, Laura Beretta, Heather L. Stevenson

## Abstract

**Background and Aims:** In clinical trials for reducing fibrosis in NASH patients, therapeutics that target macrophages have had variable results. We evaluated intrahepatic macrophages in patients with non-alcoholic steatohepatitis to determine if fibrosis influenced phenotypes and expression of CCR2 and Galectin-3.

**Approach & Results:** We used nCounter to analyze liver biopsies from well-matched patients with minimal (n=12) or advanced (n=12) fibrosis to determine which macrophage-related genes would be significantly different. Known therapy targets (e.g., CCR2 and Galectin-3) were significantly increased in patients with cirrhosis.

However, several genes (e.g., CD68, CD16, and CD14) did not show significant differences, and CD163, a marker of pro-fibrotic macrophages was significantly decreased with cirrhosis. Next, we analyzed patients with minimal (n=6) or advanced fibrosis (n=5) using approaches that preserved hepatic architecture by multiplex-staining with anti-CD68, Mac387, CD163, CD14, and CD16. Spectral data were analyzed using deep learning/artificial intelligence to determine percentages and spatial relationships. This approach showed patients with advanced fibrosis had increased CD68+, CD16+, Mac387+, CD163+, and CD16+CD163+ populations. Interaction of CD68+ and Mac387+ populations was significantly increased in patients with cirrhosis and enrichment of these same phenotypes in individuals with minimal fibrosis correlated with poor outcomes. Evaluation of a final set of patients (n=4) also showed heterogenous expression of CD163, CCR2, Galectin-3, and Mac387, and significant differences were not dependent on fibrosis stage or NAFLD activity.

**Conclusions:** Approaches that leave hepatic architecture intact, like multispectral imaging, may be paramount to developing effective treatments for NASH. In addition, understanding individual differences in patients may be required for optimal responses to macrophage-targeting therapies.

## INTRODUCTION

Fatty liver disease is increasing in incidence and is predicted to become the main etiology leading to liver transplantation for both men and women [1]. Non-alcoholic fatty liver disease (NAFLD) is the most common cause of liver disease worldwide and affects approximately 25% of the population [2]. In the United States, the prevalence of non-alcoholic steatohepatitis (NASH) is estimated to be between 1.5% to 6.45% and it is commonly associated with dyslipidemia, obesity, metabolic disease, and type 2 diabetes [3] [4]. NAFLD is defined as the presence of hepatic steatosis (at least 5%) without evidence of hepatocellular injury, while NASH is defined as the presence of hepatic steatosis (at least 5%) in the presence of hepatocyte injury, including ballooning degeneration [4]. Multiple stimuli are thought to lead to macrophage activation and the progression to NASH from NAFLD, including cholesterol metabolites, adipokines, gut-derived endotoxins, and damage-associated molecular patterns (DAMPs) from injured hepatocytes [5, 6]. Under normal conditions, Kupffer cells maintain liver homeostasis and repair tissue after injury. However, in patients with steatohepatitis, Kupffer cell activation causes the recruitment of systemic macrophages by the interaction of CCL2/CCR2, promoting inflammation, immune activation, and development of hepatic fibrosis [7]. Advanced fibrosis is the most critical histologic feature of NASH that is an independent predictor of poor clinical outcome and can lead to end-stage liver disease and hepatocellular carcinoma (HCC) [8, 9].

A recent analysis of normal liver using single-cell RNA sequencing revealed the presence of two CD68+ macrophage populations, one with inflammatory properties and increased Mac387 expression, and the other with immunoregulatory/tolerogenic functions and increased CD163 expression [10]. CD68 has been shown to primarily identify resident intrahepatic macrophages, while Mac387 has been used to identify systemic macrophages that infiltrate the liver after injury [11] in a CCL2-dependent manner [12]. CD163 identifies the prototypical “M2” or pro-fibrotic macrophage that increases in number during hepatic fibrosis progression [13]. Activated hepatic stellate cells have been shown to cause infiltration and differentiation of pro-fibrotic CD163+ macrophages via the CCL2/CCR2 pathway [13]. Circulating monocytes that can enter the liver have also been characterized as inflammatory or anti-inflammatory based on the enhanced expression of CD14 or CD16, respectively [14, 15]. However, less is known about these markers on intrahepatic macrophages, although a CD14+CD16+ dual-positive population has been shown to be associated with the progression of NASH fibrosis [16].

Therapies that target or inhibit pathogenic, pro-fibrotic macrophages are currently in clinical trials [17] (**Table S1**). Certain drugs, such as CCR2/CCR5 inhibitors (e.g., cenicriviroc), farnesoid X receptor agonists (e.g., obetacholic acid), and Galectin-3 antagonists (e.g., GR-MD-02), have shown promise in decreasing hepatic fibrosis in NASH [7, 18, 19]. For cenicriviroc, data from the CENTAUR study’s year 1 analysis showed fibrosis improvement (by > 1 fibrosis stage) without impact on steatohepatitis in ∼ 20% of patients when compared to placebo. However, continued improvement in fibrosis was not observed at the end of year 2 [7, 20, 21]. Previously, we showed that multispectral imaging can characterize numerous phenotypes of intrahepatic macrophages in patients with chronic liver diseases, including NASH [22]. For this study, liver biopsies from patients with either minimal or advanced fibrosis due to NASH were used to further study macrophages *in situ* within the hepatic microenvironment. We stained liver biopsy sections with multiplex antibody panels including anti-CD68, Mac387, CD163, CD14, and CD16 antibodies, as well as with antibodies against macrophage-directed therapeutic targets, including CCR2 and Galectin-3. Multiplex immunofluorescence images were acquired using multispectral imaging and the data were analyzed using advanced imaging analysis that included algorithms with deep learning and artificial intelligence modules. Intrahepatic macrophages were characterized to determine phenotype percentages and spatial relationships. We hypothesized that patients with cirrhosis would have an increased prevalence of infiltrating macrophages, enhanced spatial relationships and enrichment of macrophage phenotypes, and more expression of treatment targets, like CCR2. In addition, since only a small percentage (∼20%) of patients showed improvement after cenicriviroc treatment in clinical trials, we also predicted that there would be variability in the macrophage phenotype percentages and marker expression between individual patients with NASH. These results provide new insights regarding the pathogenic macrophage subsets present in patients with NASH and will facilitate the design of translational medicine approaches to emerging therapies for medical liver diseases.

## PATIENTS AND METHODS

### Patients and liver biopsies

All studies were conducted on de-identified, archived liver biopsies collected from 2006 to 2022 and the protocol was approved by the University of Texas Medical Branch Institutional Review Board (IRB # 13-0511 and 22-0223). Biopsies had previously been obtained as the standard of care by licensed radiologists via the percutaneous route using an 18-gauge core needle. At collection, tissue was immediately placed in 10% buffered-formalin and tissue blocks and slides were prepared and stored in a CAP-accredited laboratory by certified histotechnologists as previously described [22]. Archived tissue blocks were selected from patients (n = 39) with biopsy-proven NASH (minimal fibrosis: fibrosis stage 0-1/4, n = 18; advanced fibrosis: fibrosis stage 3.5-4/4, n = 17; and varied fibrosis stages 2-4/4, n = 4). Disease activity and fibrosis stages were determined by a board-certified pathologist using established criteria for NASH (i.e., NAS scores) [23]. Unstained sections were cut at 3µm from the tissue blocks immediately prior to staining.

Demographic and clinical data were collected for each patient in the study. Alanine aminotransferase (ALT), aspartate aminotransferase (AST), and alkaline phosphatase levels (ALP), total bilirubin, albumin, and platelets were determined at the time of biopsy and during follow-up chart review. Imaging studies were reviewed to evaluate for the presence of HCC or other liver lesions.

### RNA extraction and nCounter analysis

Three unstained FFPE liver biopsy sections (cut a 3µm) from patients 1-24 were used for NanoString nCounter analysis. RNA was extracted using the High Pure FFPET RNA Isolation kit (Roche, Germany), concentrated, and cleaned using the RNA clean & concentrator kit (Roche, Germany) at the MDACC Biospecimen Extraction Facility. Concentrations of the extracted RNA samples were measured with the Qubit 2.0 fluorometer (Thermofisher, MA) and the quality was determined using the RNA6000 pico assay (Agilent Technologies, Santa Clara, CA) at the MDACC Advanced Technology Genomics Core. RNA concentrations of 50-150ng were used for the hybridization reaction and the RNA expression profiling was analyzed using a custom-designed panel (like the PanCancer Immune Profiling Panel) for the nCounter Sprint profiler (NanoString Technologies, Seattle, WA). This panel comprises ∼40 housekeeping genes and 730 immuno-oncology-related targets [24]. The complete list of genes included in the Pan-Cancer Immune Profiling Panel is available at https://www.nanostring.com/products/ncounter-assays-panels/oncology/pancancer-immune-profiling/). Raw data (RCC files) were imported into the nSolver software and analyzed using the Advanced Analysis Module (NanoString Technologies), which allowed us to obtain normalized, log2-transformed data for each gene in each group and to determine differential gene expression.

### Liver biopsy fibrosis quantification

The fibrosis stage was first estimated using established criteria for NASH [23] and then we optimized an application to determine collagen proportionate area (CPA) [25] when Masson’s trichrome stains were available. Trichrome stains were digitally scanned on an Aperio ImageScope Digital slide scanner (Leica Biosystems, Wetzlar, Germany). We used the application (#10040) available in Visiopharm’s APP Center (https://visiopharm.com/app-center/) as a template to optimize the detection of the blue-stained areas (collagen) from the trichrome stains using a thresholding method. Liver capsules, large portal tracts (identified by the presence of large hepatic arteries or nerve bundles), torn tissue, and artifacts were manually removed. White spaces from steatosis were filled to be quantified as part of the total tissue area. The algorithm divides the amount of collagen by the total biopsy surface area to obtain the CPA (CPA%= [Σ collagen area in square microns/Σ total area in square microns × 100).

### Multiplex staining and imaging acquisition

Workflow included the development of a spectral library and the optimization of a macrophage markers panel (CD68, CD163, Mac387, CD14, CD16, and DAPI) in monoplex and multiplex configurations, as previously described [22] and applied it to patients 25-35. We also optimized a therapy-related panel, which includes two known targets of emerging anti-fibrotic therapies (CCR2 and Galectin-3), as well as CD163, Mac387, and DAPI. We applied the therapy-related panel to randomly select patients (36-39) recently diagnosed with NASH by liver biopsy for further analysis beyond NAS grading and fibrosis staging. Multiplex stains were either conducted manually or with a similar protocol on a Ventana Discovery Ultra automated tissue stainer (Roche Diagnostics, Indianapolis, IN).

Antibodies and optimization conditions used in the multiplex panels are shown in **Table S2**. Images were acquired using the Vectra 3 Quantitative Pathology Imaging system (Akoya Biosciences, Marlborough, MA). Regions of interest were obtained from at least 50% of the surface area of each liver biopsy. Multiplex images were acquired for cell segmentation and phenotyping using InForm software (Akoya Biosciences). Thresholds for specific markers were adjusted using images from control patients and then applied to all the images through batch analysis. The acquired imaging data were exported using two different approaches, cell_seg_data text files and multicomponent TIFFs.

### Imaging and spatial analyses

First, multi-component TIFF images were exported from InForm (Akoya Biosciences) and analyzed with Visiopharm software (Visiopharm, Hoersholm, Denmark), including custom tissue detection, nuclear detection, and multiplex phenotyping applications using thresholding and deep learning algorithms. Phenomap software (Visiopharm) was used to determine the location of phenotypes within the hepatic microenvironment and corresponding t-SNE plots were generated. The heterogeneity of the various macrophage phenotypes present, and their percentages, were determined using Visiopharm’s phenotype profile and deep learning algorithms. These allow for visual comparison of macrophage phenotypes present in the livers of the two patient groups. Batch analysis was used to directly compare the minimal fibrosis group to the advanced fibrosis group, and then a separate algorithm was used to compare differences in the individual patients within each group.

Second, cell coordinates and intensity characteristics were extracted as cell_seg_data text files and provided to the imaging Bioinformatics group at the University of Michigan. The heterogeneity and distribution of the various cell phenotypes in the study groups were determined using Uniform Manifold Approximation and Projection for Dimension Reduction (UMAP), a dimension reduction technique in the same vein as t-SNE used for visualization of high dimensional data [26]. UMAP allows visualization and comparison of high dimensional data, such as the cell profiles in different liver biopsies and disease stages, while preserving the innate local and global relationships in the data. For this study, we considered all 30 possible marker combinations of the macrophage subpopulations for plotting the UMAPs. Cells that were negative for all phenotype biomarkers of interest after applying the thresholds were excluded from the analysis. The implementation and visualization of this workflow were done using MATLAB 2020a (MathWorks Inc., Natick, Massachusetts)[27].

The spatial G-function is a nearest-neighbor-type cumulative distribution function used to quantify the mixing and/or infiltration of a cell of interest around a reference cell [26].

Mathematically, it can be expressed as follows:

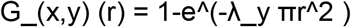

Where the subscripts ‘x’ and ‘y’ indicate that the spatial distribution of cell type ‘y’ relative to the cell type ‘x’ is being computed, ‘r’ refers to the distance from the reference cell type, and λ_y is the overall density of cell type ‘y’ on the slide.

Cell_seg files were also used for Giotto enrichment analysis. This method identifies pairs of cell types that are likely to be near one another. This is done by first using cell locations to create a k-nearest neighbor graph, where k=4. In the neighbor graph, each node represents a cell, and edges between nodes indicate “interaction” between the cells. Each cell is connected to its 3 nearest neighbors. Nodes are assigned to cell types based on the image data. To identify enriched or depleted interactions between cell type pairs, Giotto creates a random permutation distribution by shuffling cell labels on the graph. It then calculates a p-value for each cell type pair by observing how often the simulated occurrence of edges between those cell types occurred in comparison to simulations.

### Statistical analyses

Between-groups phenotype percentage analyses were performed using GraphPad Prism 9.5.0. The normal distribution of the data was determined using D’Agostino-Pearson normality test. An unpaired t-test was used to compare differences between the groups and continuous variables. The Mann-Whitney test with Holm-Sidak correction for multiple comparisons was used to compare non-normally distributed variables. The area under the curve (AUC) of this computed G-function at selected ‘r’ values was computed to characterize infiltration. Wilcoxon rank-sum test was used to identify significance in pairwise differences between groups of patients with NASH. The implementation and visualization of this workflow were done using R software [28]. Boxplot figures visualizing the differences in significant groups were plotted in R using ggplot2 [29]. A p-value < 0.05 was considered to represent statistical significance.

## RESULTS

A graphical summary of the study, approach, and main conclusions is shown in **Figure 1**. For the first set of analyses, RNA was extracted from homogenized FFPE liver biopsies that had been collected from well-matched patients with either minimal (fibrosis stage 1/4, n = 12) or advanced fibrosis (fibrosis stage 4/4, n = 12), and then analyzed using nCounter. We were primarily interested in determining differences in expression of macrophage- and therapy-related genes relevant to the development of fibrosis and those that were being targeted in recent clinical trials (e.g., CD68, CD16, CD14, CD163, CCR2, and Galectin-3) [7, 11, 13, 19, 30]. The demographics and laboratory data for these patients (#1 -24) are shown in **Table S3**. Several markers that are known to be increased in the hepatic fibrogenesis (e.g., CD68 [31] and CD16 [32]) showed no significant difference between well-matched patients with minimal or advanced fibrosis (**Figure 2B**). Of note, CCR2/CCL2/CCL5 and Galectin-3 (LGALS3), targets of cenicriviroc and galactoarabino-rhannogalaturonan/GB1211, respectively, were significantly increased in the patients with advanced fibrosis (**Figure 2B**). Macrophage genes like CD206, CD163, and ARG1, which play important roles in liver homeostasis and profibrotic functions [13] were significantly (p<0.05) downregulated in patients with advanced fibrosis (**Figure 2B**). A complete list of gene expression in matched nonalcoholic steatohepatitis patients with advanced and minimal fibrosis, included in the heatmap (**Figure 2A**) is shown in **Table S4**. These unexpected results prompted us to further evaluate macrophage phenotypes and expression of these targets *in situ* in the hepatic microenvironment with spectral imaging, an approach that preserves architecture.

**Figure 1.**
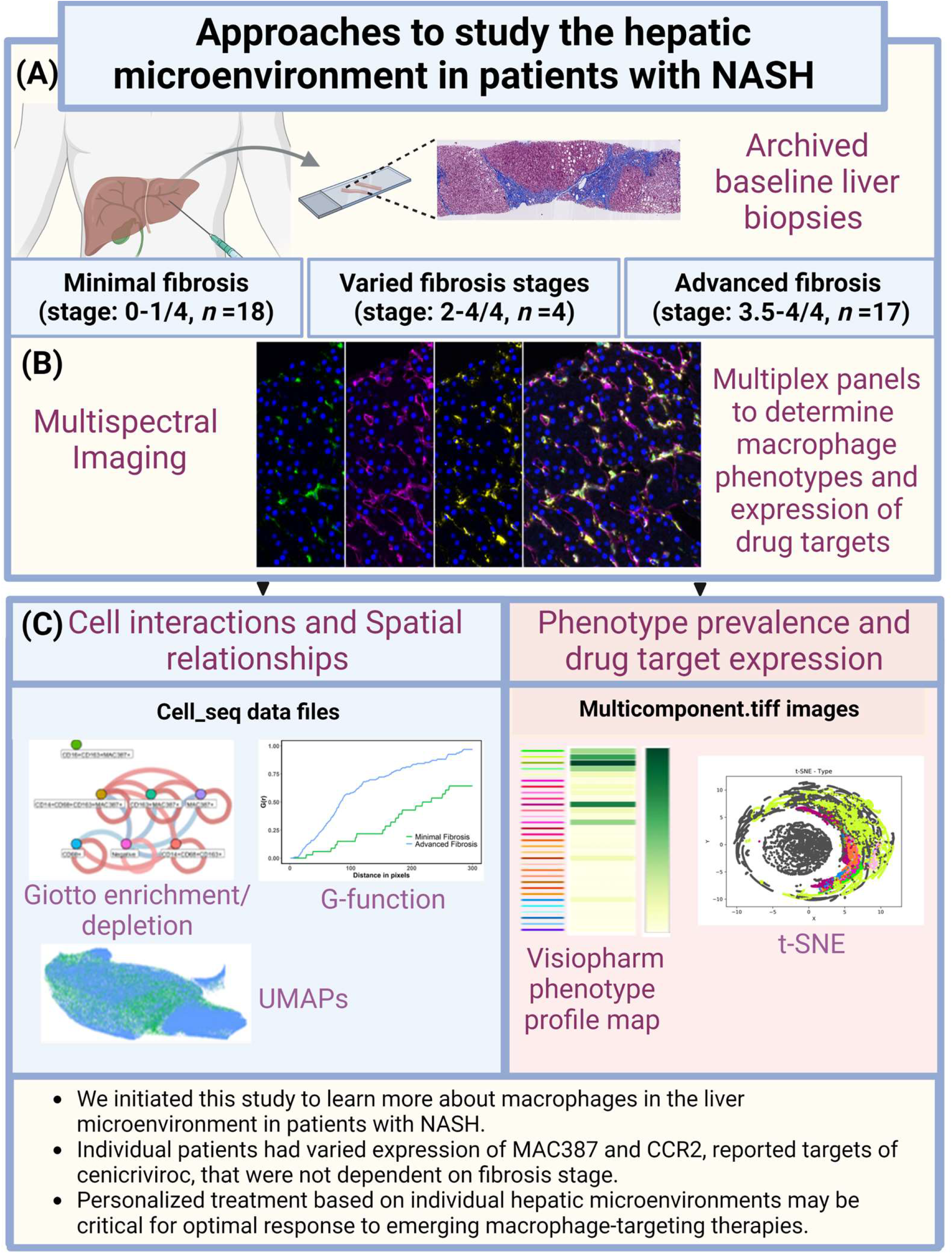
Assay development and imaging analysis workflow. (A): Liver biopsy tissue blocks were acquired from patients with either minimal (stage 0-1) or advanced fibrosis (incomplete cirrhosis or cirrhosis) due to NASH. **(B):** First, unstained slides from FFPE liver biopsy tissue blocks were used to extract RNA from patients with NASH and either minimal or advanced fibrosis. Differences in gene expression in these two groups were analyzed with the NanoString sprint platform using nCounter. **(C)**: For spectral imaging analysis, optimal conditions for antibodies, fluorophores and antigen retrieval buffers were determined [22], unstained slides were cut (at 3*μ*m) from each patient’s tissue block immediately prior to multiplex staining. After staining, slides were scanned with the Vectra 3 Automated Quantitative Pathology Imaging system (Akoya Biosciences), regions of interest were selected, and images were unmixed, segmented, and phenotyped using InForm software (Akoya Biosciences). The Phenochart application of the inForm software was used to randomly stamp multiple ROIs in each liver biopsy (totaling at least 50% of the tissue). These corresponded to 50-70 images per patient, depending on the size of the liver biopsy. Each of the ROIs were then acquired at 20X (4 representative images are shown). **(D)**: Two different approaches were used to analyze the spectral imaging data. First, Visiopharm software phenotyping modules were used to analyze multiplex images to generate phenotype maps of the tissue microenvironment, t-SNE plots, and phenotype matrix heat maps. Next, cell phenotypes were assigned according to isolated spectral signals using representative images from control patients and then applied to all samples using batch analysis. We used Uniform Manifold Approximation and Projection for Dimension Reduction (UMAP). UMAP is an algorithm for non-linear dimension reduction based on manifold learning techniques and ideas from topological data analysis that allows visualization of heterogeneity and distribution of various cell phenotypes in the study groups (MATLAB 2020a; MathWorks Inc.). Enrichment/depletion analysis was performed using Giotto, an algorithm that identifies pairs of cell types that are more or less likely to be near one another. Quantities, enrichments, and depletions of phenotypes were compared between groups of patients using a combination of Visiopharm and Giotto analyses. Created with BioRender.com

**Figure 2.**
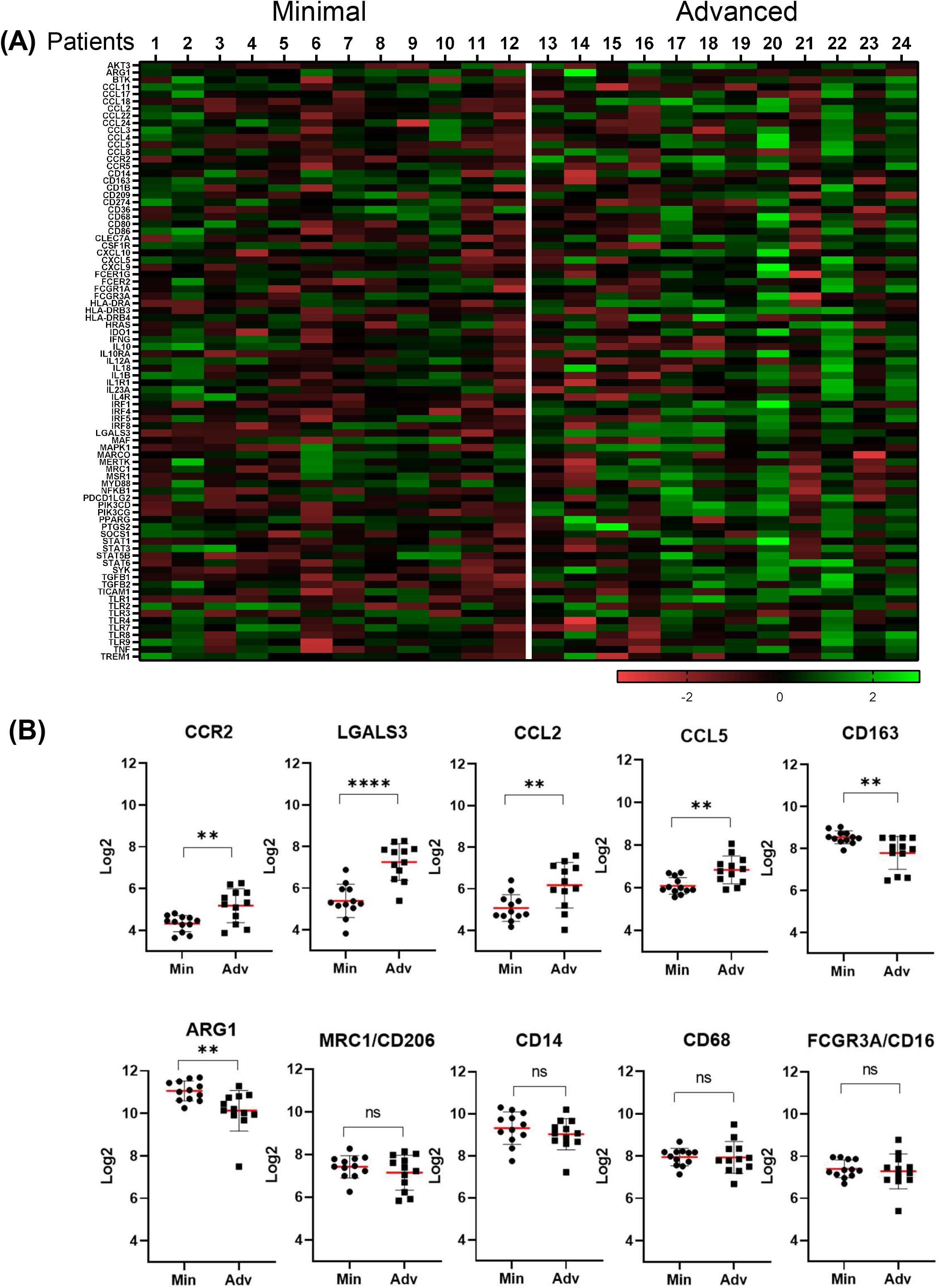
nCounter analysis showed significant differences in therapy-related markers but was unable to detect differences in several macrophage populations. Three unstained FFPE liver biopsy sections from matched patients (1-24) were used for nCounter gene expression analysis, which comprises ∼ 40 housekeeping genes and 730 immuno-oncology-related targets [24]. **(A)** Heat map displaying differential gene expression profiles of liver biopsies from NASH patients with minimal (n = 12) and advanced (n = 12) fibrosis. **(B)**. Gene expression analysis showing significant upregulation of CCR2, CCL2, CCL5 and Galectin-3 (LGALS3) and downregulation of CD206/MRC1, CD163, and ARG1, in patients with advanced fibrosis. Other genes like CD68, CD14, and CD16 were not significantly upregulated in patients with minimal or advanced fibrosis (**Figure 2C, Table S4**). An unpaired t-test was used to compare differences between the groups. *p < 0.05; **p < 0.01; ***p < 0.001; ****p < 0.0001. Min = minimal fibrosis, Adv = Advanced fibrosis.

We obtained archived liver biopsies from another group of patients with biopsy-proven NASH and again separated these into groups with either minimal fibrosis (stage: 0-1/4; n = 6) or advanced fibrosis (stage: 3.5-4/4; n = 5). Demographic information, laboratory data, and histopathologic scoring determined at the time of baseline biopsy from the groups of patients (#25-35) with minimal and advanced fibrosis due to NASH are shown in **Table S5**. Representative images of the trichrome stains and corresponding CPA fibrosis quantification from each of the patient groups (A: patient 25 with minimal fibrosis; B: patient 31 with advanced fibrosis) is shown in **Suppl Figure 1**. The fibrosis stage was determined in two ways. It was first estimated using the NASH criteria [23] and then more objectively measured using the CPA [25]. The calculated CPA in the study patients showed percentages of fibrosis from 0.2 to 6.1% and 6.9 to 25.9%, in the minimal and advanced fibrosis patients, respectively, and was significantly increased (p < 0.04) in the advanced fibrosis group (**Figure S1C**). The fibrosis stages ranged from 1 to 4 (out of 4) (**Table S5**). Representative multiplex immunofluorescence images from patients with either minimal (patient 25) or advanced (patient 34) fibrosis are shown in **Figure 3A and 3D**, respectively. From each of these 5-color multiplex immunofluorescence images, a Visiopharm phenotyping module was used to display corresponding phenotype maps where each colored dot represents a unique cellular phenotype (**Figure 3B and 3E**). Patients with advanced fibrosis had more inflammation in the portal tracts and CD68+ cells were more numerous. Infiltrating systemic macrophages were observed in the portal tracts of both groups (see Mac387+), although they were more frequent in the patients with advanced fibrosis. t-SNE plots were generated from batch analysis of all the images from each patient group, showing different patterns of phenotype distribution between the groups (**Figure 3C and 3F**). More phenotypes were identified in the phenotype profiles (cutoff: 0.1%) generated from the patients with advanced fibrosis when compared to those with minimal fibrosis (**See Fig 3G**). The patients with minimal fibrosis had more CD14+ on cells lining the hepatic sinusoids (see **Figure 3A, magenta**) and the corresponding t-SNE plot showed larger spread of CD14+ cells when compared to patients with advanced fibrosis (compare **Figure 3C with 3F; 3G, light green**). CD68+, CD16+, Mac387+, and CD163+ cells were more frequent in the patients with advanced fibrosis (compare **Figure 3F with 3C; 3G**). To compare differences in the prevalence of phenotypes between the two groups of patients, we compared the percentages of each of the observed phenotypes from the Visiopharm image analysis (**Figure 3G**). Our Visiopharm workflow combines deep learning and artificial intelligence to allow for accurate cell segmentation and multiplex phenotyping of tissue microenvironments using all the multiplex images acquired from each group. We identified 30 different phenotypes from the multiplex spectral images that had a prevalence greater than zero. Patients with advanced fibrosis had an increased prevalence (>3 folds) of CD68+, CD16+, CD163+, CD16+CD163+, and CD68+CD16+CD163+ macrophages in the hepatic microenvironment when compared to patients with minimal fibrosis. However, no significant differences were observed between these two groups. Several populations showed similar prevalence (< 1-fold) in both groups of patients with NASH, including CD14+CD16+, CD14+CD16+CD163+, and CD68+CD14+CD16+CD163+. CD14 expression thought similar between the groups, was the only one that slightly lower (<1 fold) in patients with advanced fibrosis, like in a previous study [22].

**Figure 3.**
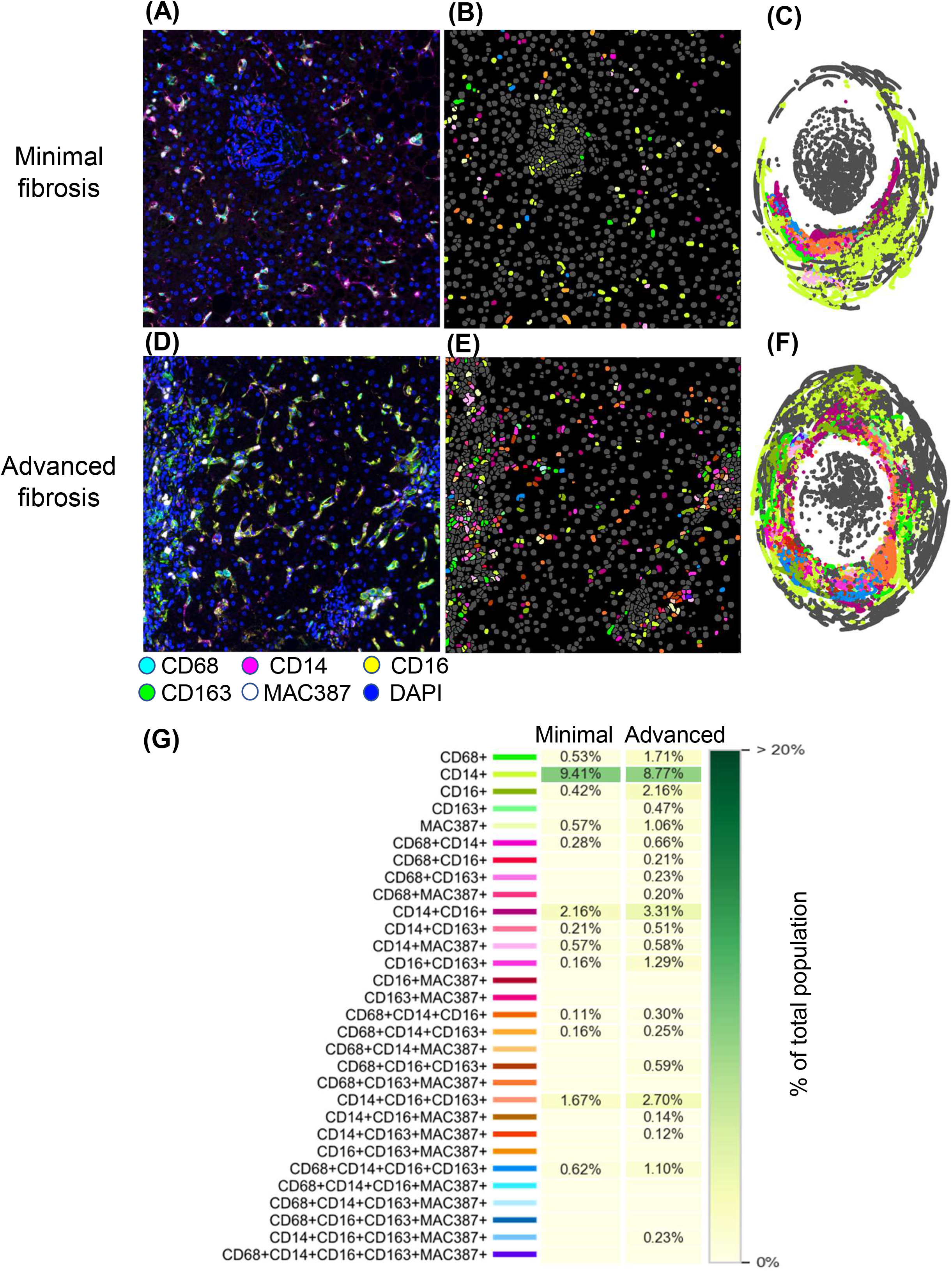
Phenotypic maps, t-SNE cluster analysis, and phenotype profile plots, highlighted unique cell populations in patients with advanced fibrosis due to NASH. (A, D): A single unstained liver biopsy slide from each patient (minimal fibrosis: patients 25-30; advanced fibrosis: patients 31-35) was stained with the multiplex immunofluorescent panel (CD68: Opal 520, CD14: Opal 540, CD163: Opal 650, CD16: Opal 620, Mac387: Opal 690, and nuclear stain: DAPI). ROIs (totaling at least 50% of each liver biopsy) were collected from each patient and representative images from each group are shown for comparison. **(A)**: Shows a representative patient with minimal fibrosis (patient 25) due to NASH. Patients with minimal fibrosis had fewer CD68+ and CD16+ cells and increased expression of CD14+ within the sinusoids. **(D)**: NASH patients with advanced fibrosis (patient 34) had more macrophages (CD68+, Mac387+ and CD163+) present within both the portal tracts and lobules. **(B-C, E-F)**: Multicomponent TIFF images from inForm were uploaded into Visiopharm for analysis using custom-designed algorithms for analysis. First, phenomaps were generated that identified and characterized unique phenotypes by labelling each with a different color dot **(B-E)**. Cells that did not stain with any of the multiplex antibodies appear gray (e.g., hepatocytes). **(C, F)**: Next, t-SNE plots were generated, allowing unique cell clusters to be compared between the different groups. The colors of phenotypes in the phenomaps and the t-SNE plots correlate with the phenotype colors shown in Figure 3G. **(G)**: To compare the prevalence of the different phenotypes between each group, we used a customized phenotype profile script from Visiopharm. Patients with advanced fibrosis due to NASH had increased percentages of CD68+, CD16+, and CD163+ macrophages when compared to the minimal fibrosis group. Both groups of patients with NASH had Mac387+ macrophages. A CD16+/CD163+ dual-positive population was more prevalent in patients with advanced fibrosis. Mann-Whitney test followed by Holm-Sidak correction for multiple comparisons was used to compare differences between the groups. No significant differences were observed between the minimal and advanced fibrosis groups. Phenotype profile cutoff: 0.1%.

Next, we evaluated the NASH patients individually to determine if heterogeneity existed between the patients in each group (**Figure 4)**. Most of the patients with advanced fibrosis had an increased prevalence of CD68+ when compared to patients with minimal fibrosis, except for patent 32. Patient 29 in the minimal fibrosis group had a higher prevalence of CD68+ macrophages when compared to the rest of the patients in this group. The prevalence of systemic Mac387+ macrophages was heterogeneous in both groups, with higher prevalence observed in a couple of patients within each group (**Figure 4**). In the minimal fibrosis group, patients 27 and 29 had greater percentages of Mac387+ macrophages when compared to the other patients. In the advanced fibrosis group, patient 32 had the highest percentage of CD14+ expression. In contrast, patient 34 had the greatest number of unique phenotypes in the hepatic microenvironment and many of these were not present in other patients in this group. Patients 33 and 34 in the advanced fibrosis group had the highest percentage of Mac387+ macrophages, while in patient 32 this population was the lowest. In the advanced fibrosis group, several of the patients had variable numbers of the different phenotypes including CD68+, Mac387+, CD16+CD163+, and CD14+CD16+CD163+.

**Figure 4.**
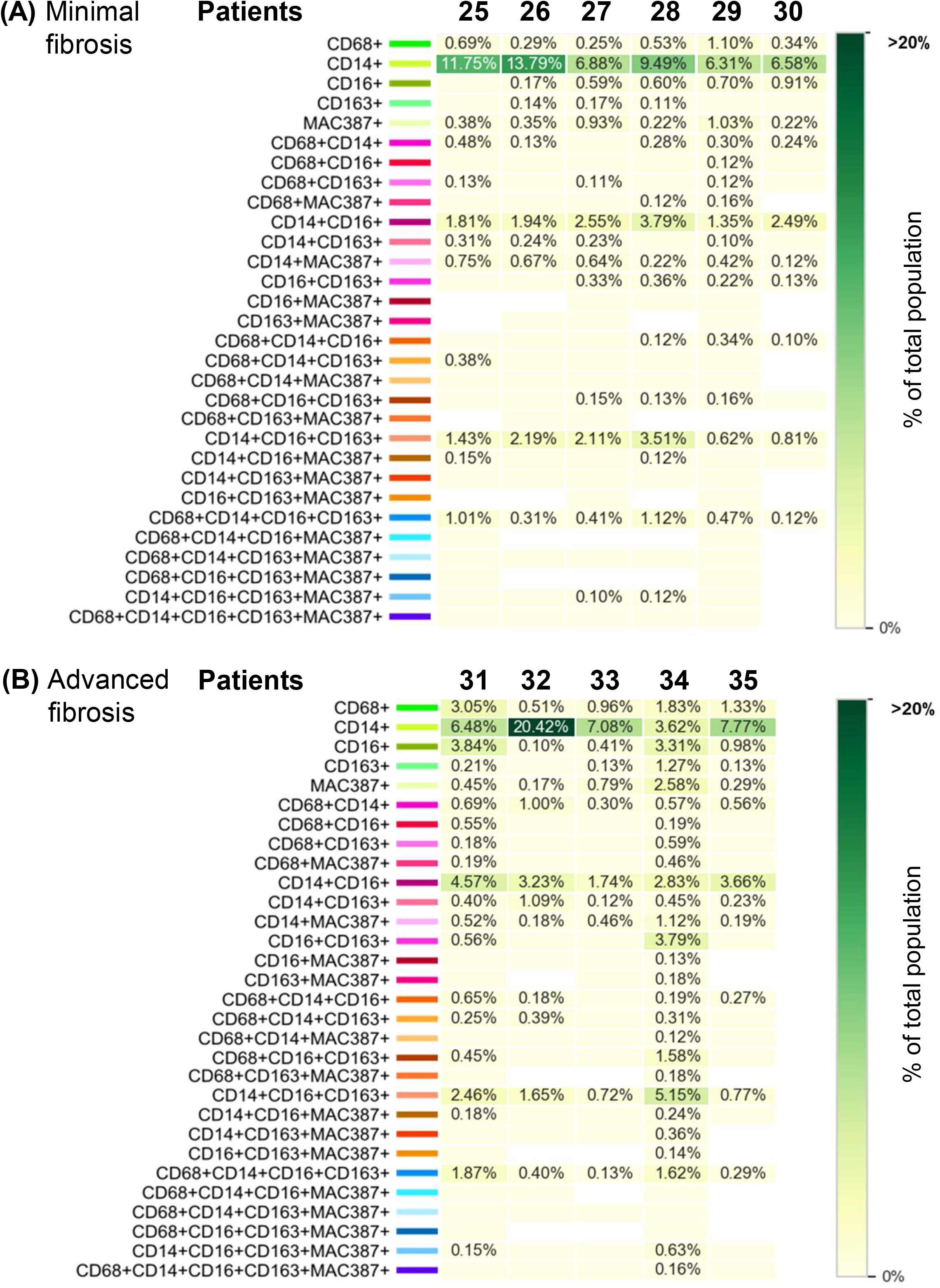
Individual patients with NASH have variation in intrahepatic macrophage populations. (A-B): We used a custom-designed Visiopharm algorithm to determine the prevalence of the phenotypes identified in Figure 3G in each of the individual patients that had minimal (**A**, patients 25-30) or advanced fibrosis (**B**, patients 31-35) due to NASH. Within each group, certain patients had increased marker expression or populations when compared to others. CD68+ had increased percentages in most of the patients with advanced fibrosis and was also increased in patient 29 from the minimal fibrosis group. Mac387+ expression was variable in both groups of patients. A dual positive, CD14+CD16+, population was identified in both groups of patients, although there was variable prevalence among individual patients. A CD14+CD16+CD163+ population was also identified in all the patients. Patient 34 had high percentages of several phenotypes, several of which were not identified in the other patients. Patients with advanced fibrosis had increased percentages of phenotypes that were absent in most patients with minimal fibrosis. Mann-Whitney test followed by Holm-Sidak correction for multiple comparisons was used to compare differences between the groups. No significant differences were observed between the minimal and advanced fibrosis groups.

In addition to comparing the unique phenotypes and phenotype percentages between the minimal and advanced fibrosis groups, we also evaluated the spatial relationships of the various phenotypes detected in the hepatic microenvironment. First, we utilized UMAP, a dimension reduction tool used to capture and visualize large, high-dimensional datasets and their local and global variations in cell biomarker expression (**Figure 5A-E**). UMAPs combined the data from all images in each group while preserving the innate local and global relationships in the data. The UMAP plots show the differences between these populations in patients with NASH with either minimal (green) or advanced (blue) fibrosis (**Figure 5A-C**). Cells that were negative for all phenotype biomarkers of interest after applying the thresholds were excluded from the analysis. Differential distribution of CD68+ and Mac387+ macrophages was observed in patients with advanced fibrosis (**Figure 5D-E**). **Figure 5F** shows a representative G-function curve, where a steeper slope indicates more infiltration of the cell of interest near the reference phenotype (i.e., Mac387+ cells with CD68+ cells). **Figure 5G** compares the AUC of the G-function curves for the Mac387+ versus CD68+ phenotype pair in the groups of patients with either minimal or advanced fibrosis due to NASH. There was a significant difference (p < 0.01) in the infiltration curves between the groups, with a bigger AUC value indicative of increased infiltration of MAC387+ at smaller distances from CD68+. None of the other phenotype interaction pairs showed significance when compared using G-function analysis.

**Figure 5.**
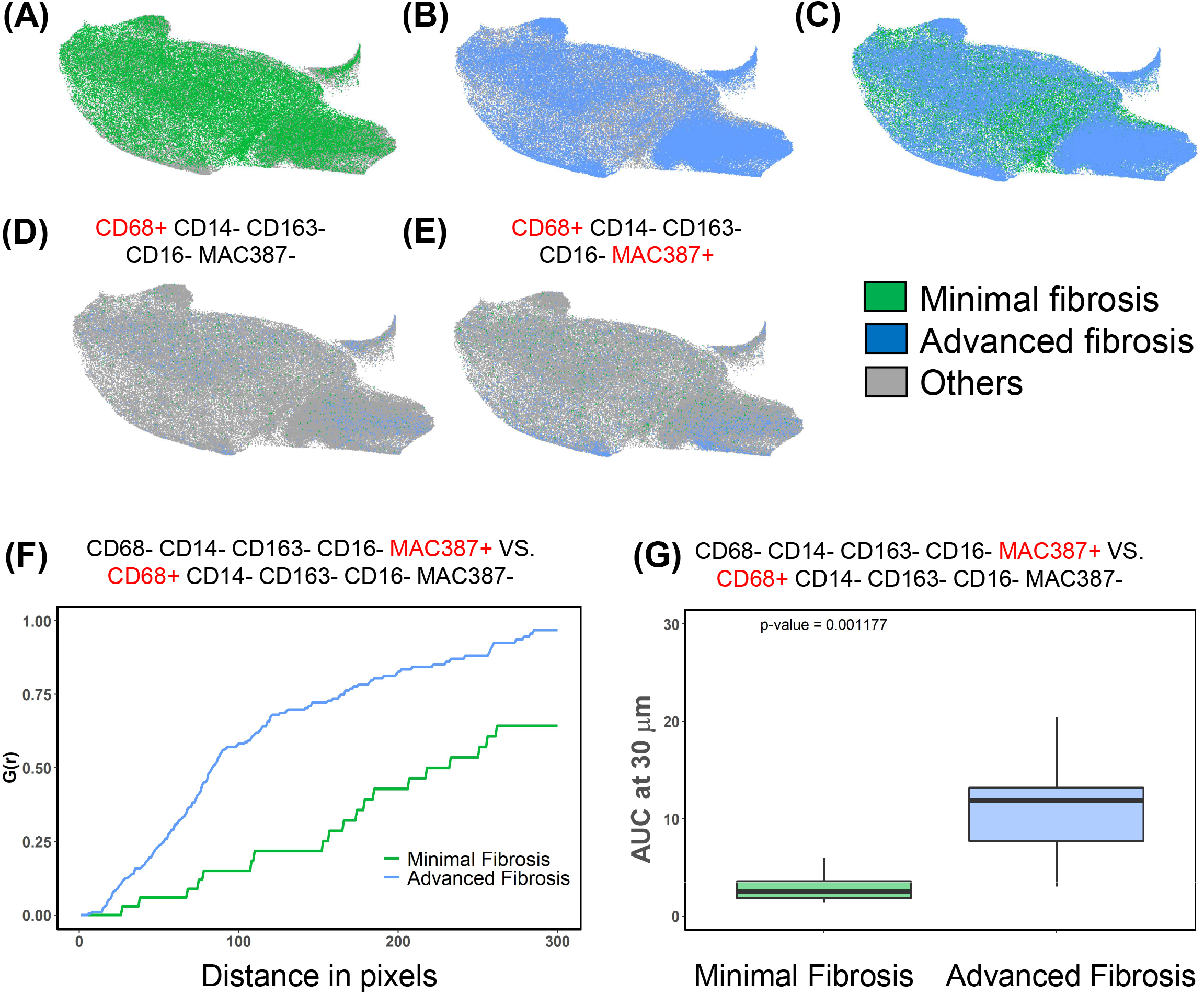
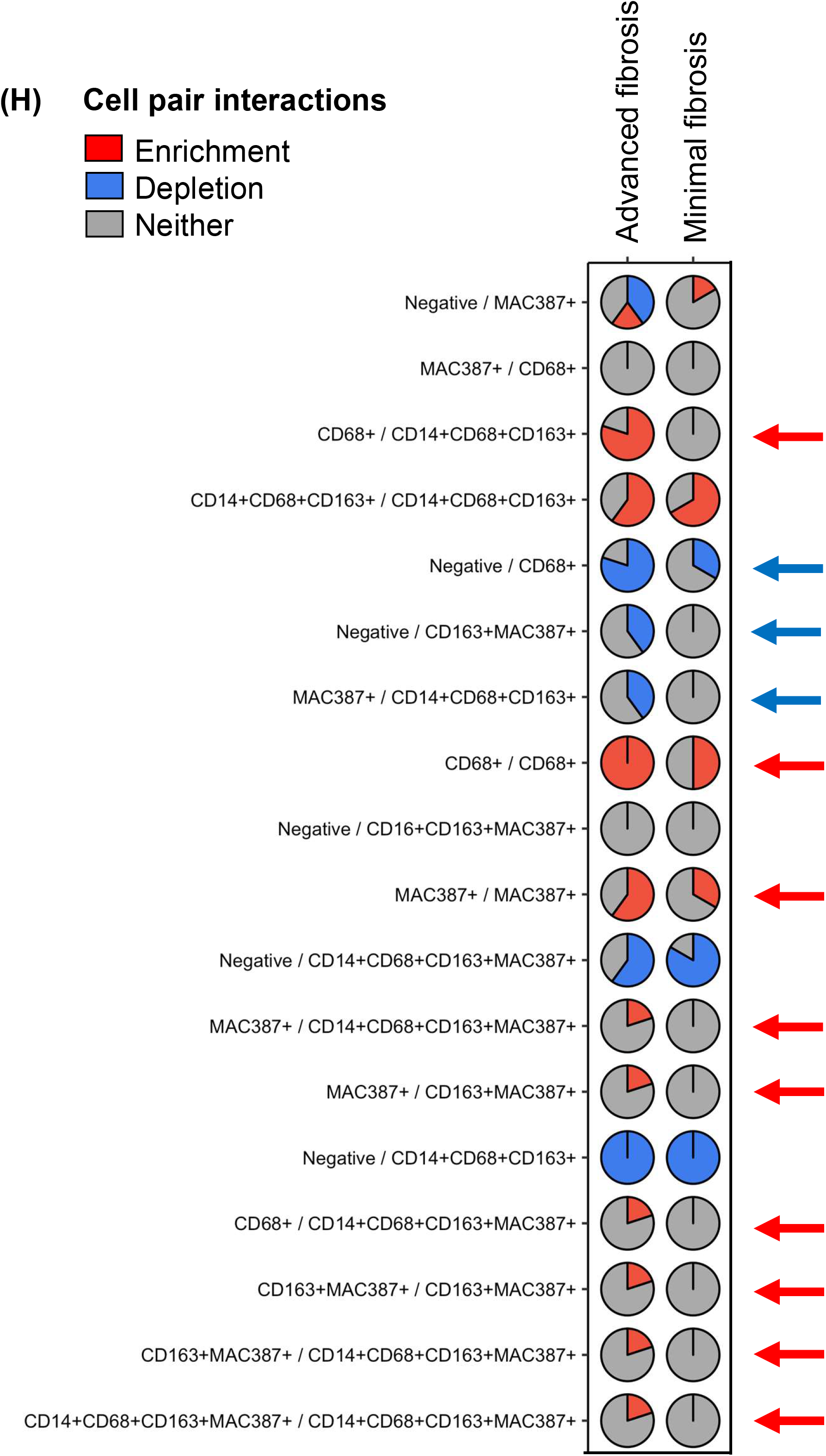
UMAPs and G-function nearest neighbor spatial analyses showed differences in infiltration between pairs of cell phenotypes in patients with minimal and advanced fibrosis. (A-E): UMAP, a dimension reduction tool used to capture and visualize large, high-dimensional data sets was used to compare the groups of patients with either minimal (green, **A**) or advanced (blue, **B**) fibrosis due to NASH. Merged **(C)**. These cluster plots showed local and global variations in cell biomarker expression across datasets and were also used to visualize the relationships of CD68+ **(D)** and Mac387+ **(E)** macrophages within the cluster analyses. **(F)**: The AUC (Area Under Curve) metric of the G-function was used as a surrogate for estimating infiltration [53]. The G-function gives the probability of having at least one cell of Class 2 within the R-pixel distance of the cell of Class 1, where R=distance. The AUC of the G-function is computed at r = 30 microns and statistical comparisons were made using a Kruskal-Wallis test. Two populations showed significant differences (CD68+ and Mac387+) and these were compared in the bar graph; a Kruskal-Wallis test showed an overall p-value of 0.0012. **(G)**: Representative AUC G-function curves for the Mac387+ versus CD68+ phenotype pair is shown. The G-function curve is interpreted in the following manner: The steeper the slope, the more the infiltration of the cell of interest near the reference phenotype. The patients with advanced fibrosis showed a marked difference in the infiltration curves, with a bigger AUC indicative of increased infiltration at smaller distances from the cells of interest. The curves represent the G-function of one patient per group. **(H)**: Enrichment analysis using Giotto identifies pairs of cell types that are more or less likely to be near one another. This is done by first using cell locations to create a k-nearest neighbor graph, where k=4. In the neighbor graph, each node represents a cell, and edges between nodes indicate “interaction” between the cells. Each cell is connected to its 3 nearest neighbors. Nodes are assigned to cell types based on the image data. To identify enriched or depleted interactions between cell type pairs, Giotto creates a random permutation distribution by shuffling cell labels on the graph. It then calculates a p-value for each cell-type pair by observing how often the simulated occurrence of edges between those cell types occurred in comparison to simulations. Using X, Y coordinates of cell centroids and prior information about cell phenotypes, the Giotto package creates a nearest neighbor graph. Then, the cell labels for the graph are shuffled to create a null distribution. Each row represents one cell-pair interaction. For each cell-pair interaction, the proportion of patients in each diagnosis with an enrichment, depletion, or no significant relationship is represented by a pie chart. Rows are ordered based on how much the distribution of enrichment, depletion, and no significance proportions differs across the two diagnosis groups. The y-axis shows each of the phenotypes detected and the x-axis (at the top) shows the different patient groups. Several enrichment interactions were increased in patients with advanced fibrosis due to NASH, when compared to those with minimal fibrosis. Interactions of CD68+ macrophages with other CD68+ cells and Mac387+ macrophages with other Mac387+ cells, were enriched in patients with advanced fibrosis (red arrows). Several of the CD68+ or CD163+ populations that were proinflammatory (i.e., also CD14+) showed increased enrichment interactions in advanced fibrosis. Finally, in patients with advanced fibrosis due to NASH several enrichment interactions were observed that were not present in any of the other groups, and every one of these cell-pair relationships included increased interactions between Mac387+ cells, often with other Mac387+ populations. Interaction type: red = enrichment; blue = depletion; gray = neither.

In addition to spatial analyses using the G function, we also compared the interaction of phenotypes within the hepatic microenvironment using Giotto enrichment analysis. Using X, Y coordinates of cell centroids and prior information about cell phenotypes, the Giotto package creates a nearest neighbor graph in each of the different groups using a custom-designed algorithm that allowed the generation of a spatial analysis matrix plot (**Figure 5H**). The y-axis shows each of the phenotype interactions detected and the x-axis (at the top) shows the different patient groups. Cell-pair enrichment interactions are shown in red, while cell-pair depletion interactions are shown in blue. Several enrichments were increased in patients with advanced fibrosis and these interactions often included CD68+ and Mac387+ (**Figure 5H**, red arrows) as well as the proinflammatory macrophage marker, CD14. Three depletion interactions were identified in patients with advanced fibrosis when compared to those with minimal fibrosis (**Figure 5H**, blue arrows). Even though patients with both minimal and advanced fibrosis had a similar prevalence of Mac387+ macrophages in their livers, patients with advanced fibrosis had enhanced spatial interactions and enrichment of this phenotype when compared to patients with minimal fibrosis.

Since we observed heterogeneity in the prevalence of different phenotypes among the various patient groups, we wanted to determine if certain patients also had unique enrichment or depletion interactions and evaluated each patient on an individual basis using Giotto enrichment analysis. Giotto enrichment/depletion analysis identifies pairs of cell types that are more or less likely to be near one another. In **Figures 6A-D**, each of the colored dots represents a different macrophage phenotype and the red or blue lines indicate cell-pair interactions. The phenotypes that are compared are shown in the colored figure legends. Red lines indicate cell-pair interactions that are enriched, while blue lines indicate cell-pair interactions that are depleted.

**Figure 6.**
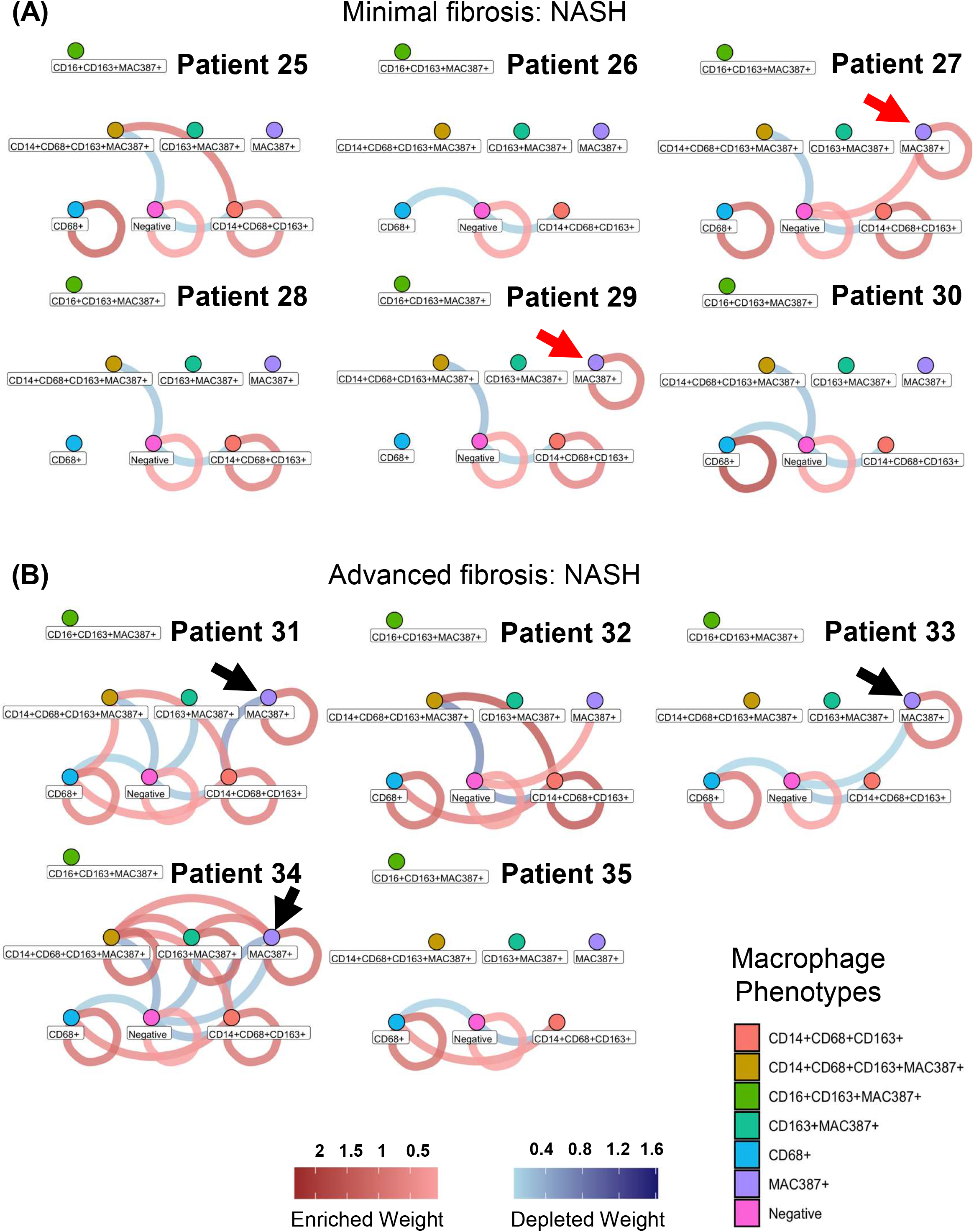
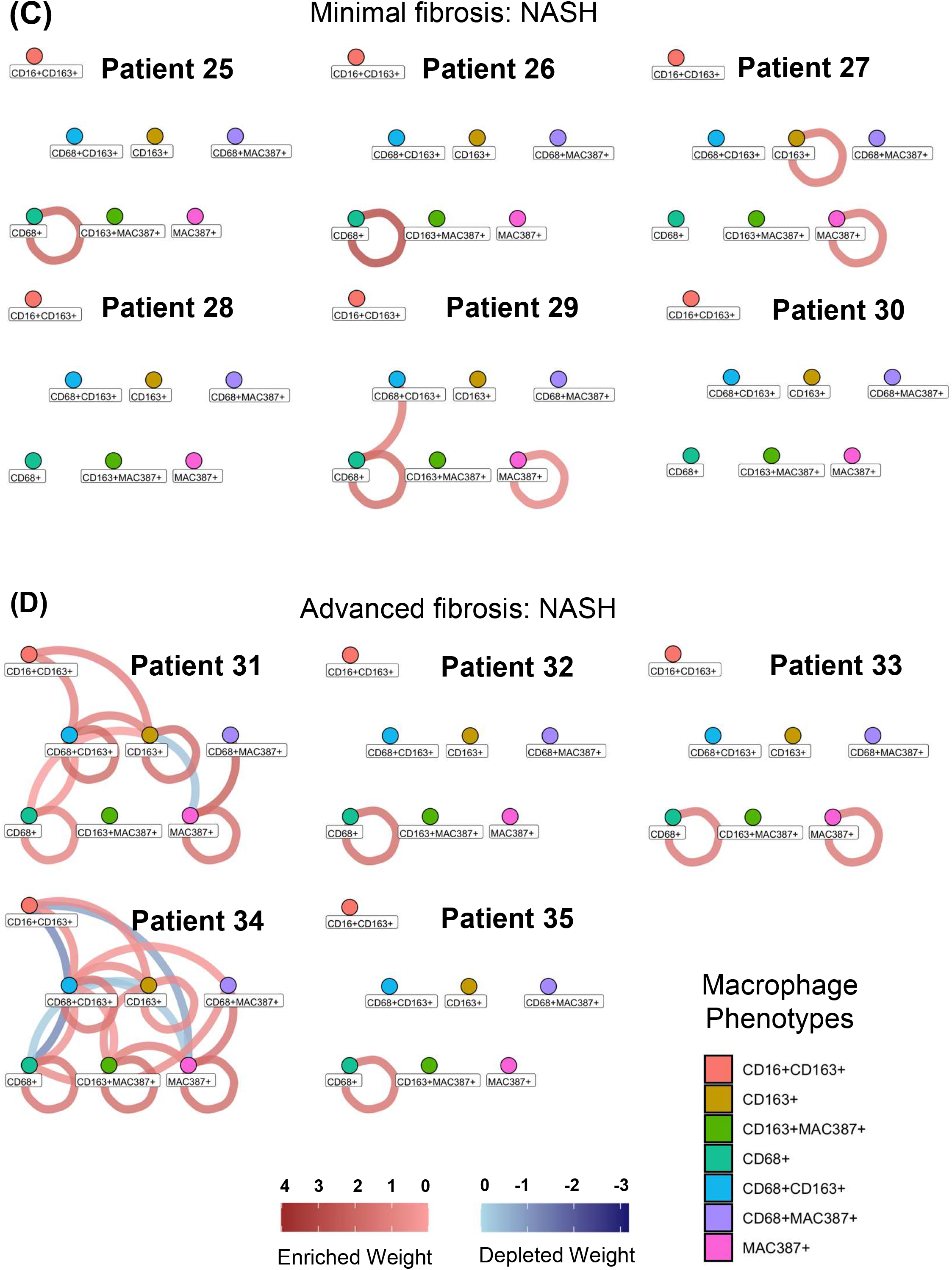
Individual nearest neighbor spatial analysis highlighted variations among individual patients with minimal and advanced fibrosis. (A-D): Individual patients in each group were analyzed separately to determine if variations existed among different patients with NASH. Each network represents the cell-pair interaction network of a single patient. For each network, a node represents a cell type. When an edge exists between a pair of nodes, this indicates a significant relationship exists between these two cell types for this patient. If two cell types share a red edge, they have an enrichment relationship, meaning the two cells are “attracted” to one another more often than random. If two cell types share a blue edge, they have a depletion relationship, meaning the two cells are “repelled” by one another more often than random. If no edge exists between a cell pair, then they do not have a relationship that differs significantly from random. The brightness of an edge indicates relationship strength; dark edges indicate strong enrichment or depletion, while light edges indicate weaker enrichment or depletion. **(A-B)**: Shows individual enrichment or depletion interactions between NASH patients with either minimal **(A)** or advanced fibrosis **(B)**. Patient 27 had more enrichment interactions than any of the other patients in the minimal fibrosis group. Only patients 27 and 29 had enrichments of Mac387+ macrophages with Mac387+ macrophages (red arrows); an interaction that was also present in several of the patients with advanced fibrosis due to NASH (black arrows). Patients 31, 32, and 34 had more enrichment interactions between cell phenotypes than the other patients in this group; patient 34 had the most within this group with 9 interactions. **(C-D):** Another set of phenotype interactions also showed that patients 27 and 29 had more enrichment of Mac387+ macrophages. In this comparison of phenotypes, patients 31 and 34 showed substantially more enrichment interactions than the other patients within this group.

Individual patients in both the minimal and advanced fibrosis groups had varied macrophage interactions. In the first set of phenotype comparisons (**Figure 6A, B**), patients 27 and 29 were the only patients with minimal fibrosis that had enrichment of Mac387+ cells with Mac387+ cells (red arrows). In the advanced fibrosis group, patients 31 and 34 had more enrichment interactions than the group of patients with minimal fibrosis and the remaining patients in this group. Overall, patients with advanced fibrosis had more enrichment interactions, particularly patients 31, 32, and 34, who had several phenotypes enriched and more than other patients within this group, while patients 31, 33, and 34 had enrichment of Mac387+ (black arrows). In the second set of phenotype comparisons (**Figure 6C, D**), patients 27 and 29 again showed enrichment of Mac387+ with Mac387+ macrophages. Patient 27 also had an enrichment of CD163+ with other CD163+ phenotypes, while patient 29 had an increased enrichment of CD68+ cells with other CD68 phenotypes. In the group with advanced fibrosis, patients 31 and 34 had the most enrichment interactions, more than any of the other patients, while patients 32 and 35 showed minimal interaction in these phenotype comparisons.

Follow-up data for patients 25-35 are shown in **Table S6**. Patients 27 and 29 that had minimal fibrosis in their baseline biopsies developed evidence of portal hypertension including splenomegaly, thrombocytopenia, and elevated partial thromboplastin time approximately 7.6 and 9.4 years later, respectively. Two of the patients with advanced fibrosis died approximately three and four years later, patient 32 from end-stage liver disease, and patient 34 from “bowel rupture.” Patient 31 had compensated cirrhosis with features of mild portal hypertension. None of the patients had been diagnosed with HCC at the time of follow-up.

Results showed that even though significant differences were observed in patients with advanced fibrosis when compared to those with minimal fibrosis, individual patients had substantial variability, even in groups that were well-matched with similar fibrosis stages at the time of their baseline biopsies. Since we observed variability in several of the macrophage phenotypes that are known targets of emerging anti-fibrotic therapies in NASH (e.g., Mac387), we hypothesized that these patients may also have a varied expression of several therapeutic targets that can be detected in liver biopsies. We randomly selected the next four liver biopsies (varied fibrosis stages 2-4/4, n = 4) that were collected at our institution with confirmed NASH (one of these was from a patient that had both HCV and NASH), and stained them with a multiplex panel containing CD163, Galectin-3, CCR2, and Mac387 (**Figure 7**). The demographics and laboratory data for these additional patients (#36-39) are shown in **Table S7**. CCR2 and Galectin-3 are two of the known targets of therapeutics that are currently in clinical trials for the treatment of NASH-mediated fibrosis (**Table S1**). Representative multiplex images taken from both the portal tracts and lobules of each of these patients are shown in **Figure 7 (A: portal tracts; B: lobules)**. Varied expression of CCR2 and Galectin-3 was detected in patients with NASH, including in patients with similar fibrosis stages (compare patient 36 with patient 37 and patient 38 with 39). CCR2 expression was not only associated with Mac387+ macrophages that were present within the portal tracts but also with the CD163+ population, while Galectin-3 was observed in both the portal tracts and lobules on both Mac387+ and CD163+ macrophages. The patient that had HCV in combination with NASH had the highest expression of CCR2 and the most advanced fibrosis in the group (**Figure 7, patient 39**). **Figure 8** shows heat maps and bar graphs to compare the expression of CD163, CCR2, Galectin-3, and Mac387 between these four additional patients. Differences were observed between the portal tract and lobular regions (**compare Figure 7A to 7B and 8A to 8B**). However, patients 37 and 39 still had the highest expression of CD163, CCR2, and Galectin-3 regardless of the region analyzed, which was also confirmed with the images from the portal tract and lobular regions combined (**Figure 8C**). The bar graphs highlight the statistical differences observed for each of the markers, in portal tracts and lobules, in the four patients (**Figure 8D-F**). Analysis of the therapeutic panel with an optimized Visiopharm algorithm identified 15 different phenotypes (**Figure 8G)** in this set of patients with NASH. CCR2+ and Galectin-3+ were more frequently expressed on, or near to, CD163+ macrophages than on Mac387+ cells (**Figure 8G**). These results indicate that individual patients with NASH not only have variable prevalence of macrophages in their livers but also unique spatial interactions with a varied expression of potential therapeutic targets.

**Figure 7.**
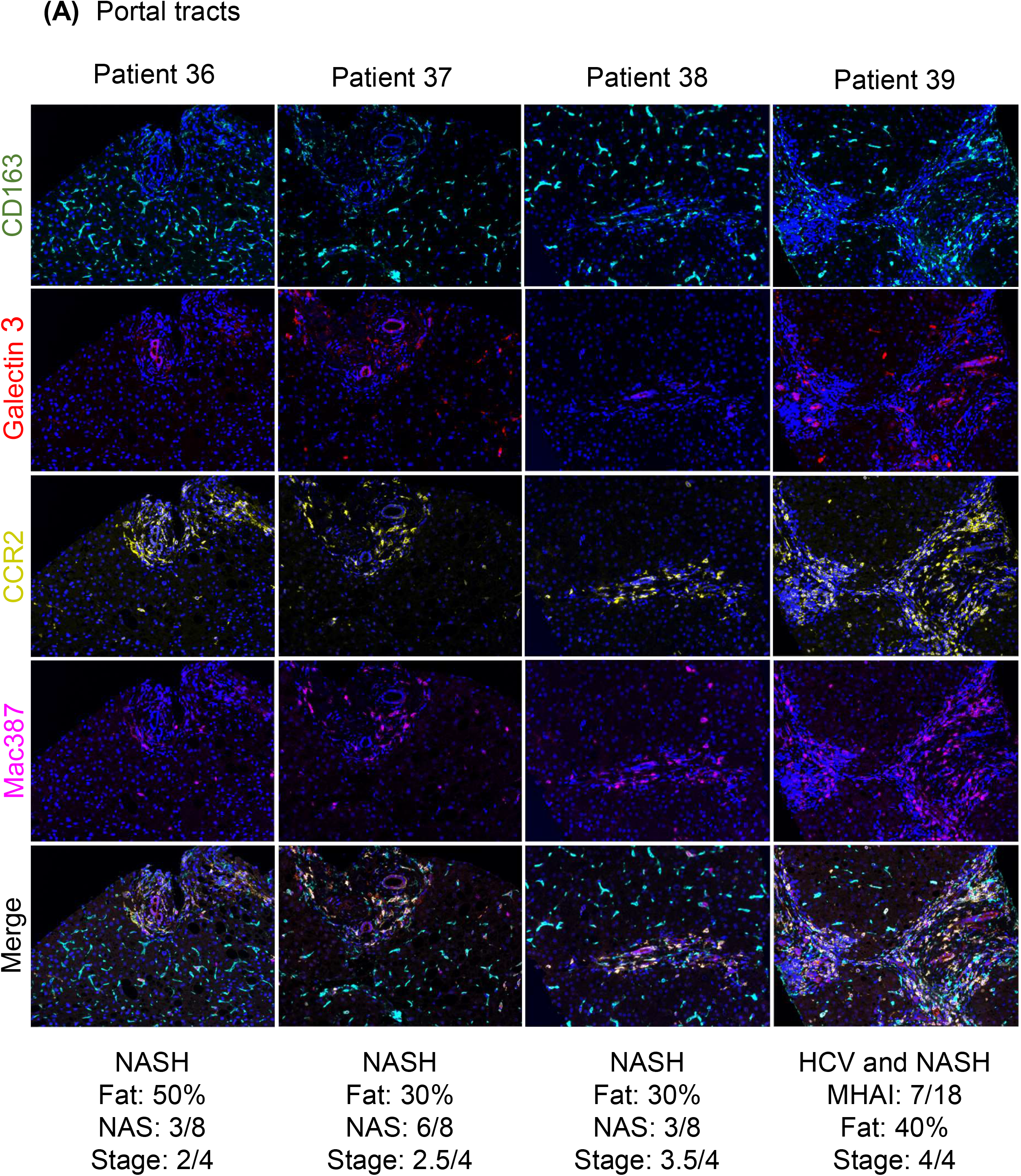
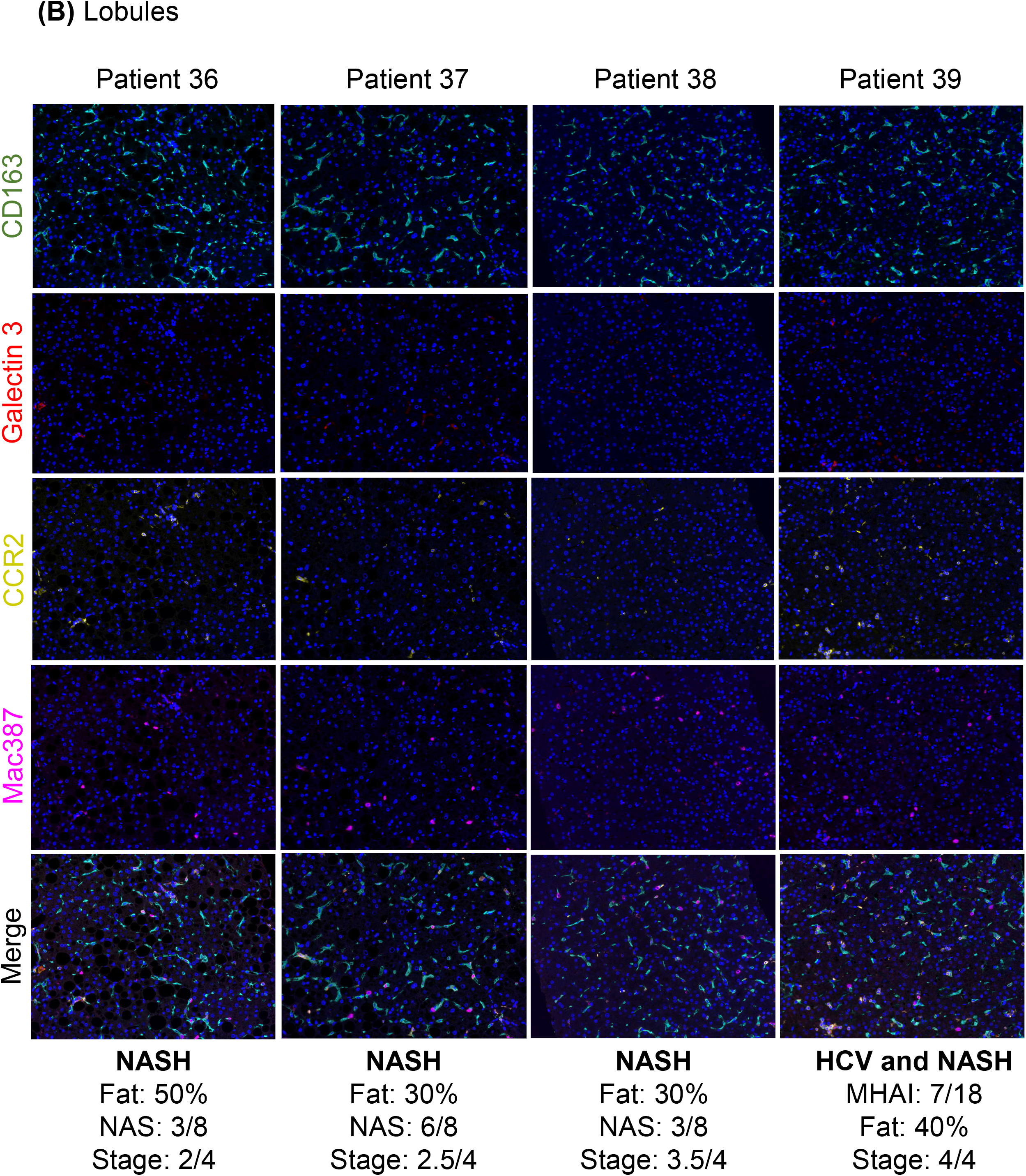
Comparison of individual patients with NASH showed heterogeneity in intrahepatic macrophages and variable expression of therapeutic targets. (A, B): We compared liver biopsies from several patients (patients 36-39) with different fibrosis stages (patient 36: 2/4, patient 37: 2.5/4, patient 38: 3.5/4, and patient 39: 4/4) due to NASH and cirrhosis due to combined HCV/NASH (patient 15). Liver biopsy slides from each patient were stained with the multiplex immunofluorescent panel (CD163: Opal 520, Galectin-3: Opal 540, CCR2: Opal 650, MAC387: Opal 690, and nuclear stain: DAPI). Representative images, collected from at least 10 portal tract regions **(A)** and 10 lobular regions **(B)**, are shown. Expression of each marker in individual patients was varied, even in those with similar fibrosis stages (e.g., patients 37 and 38). Patients 37 and 39 had the most expression of Galectin-3. The patient with cirrhosis due to HCV and NASH (patient 39) had more expression of CCR2, while one of the patients with moderate fibrosis (patient 37) had the highest expression of CD163 and Galectin-3, although CCR2 was also highly expressed in this patient. MHAI: modified hepatic activity index.

**Figure 8.**
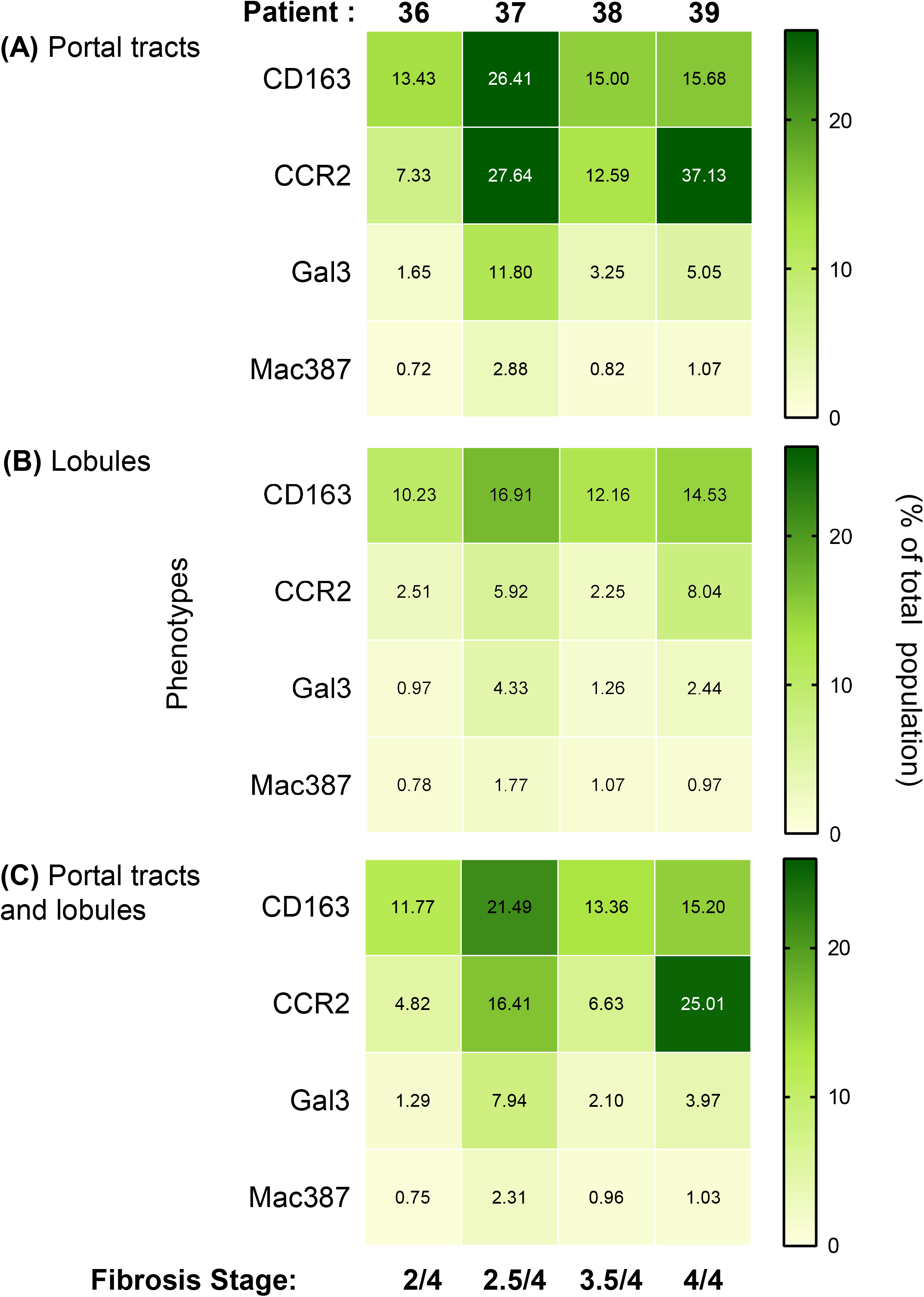
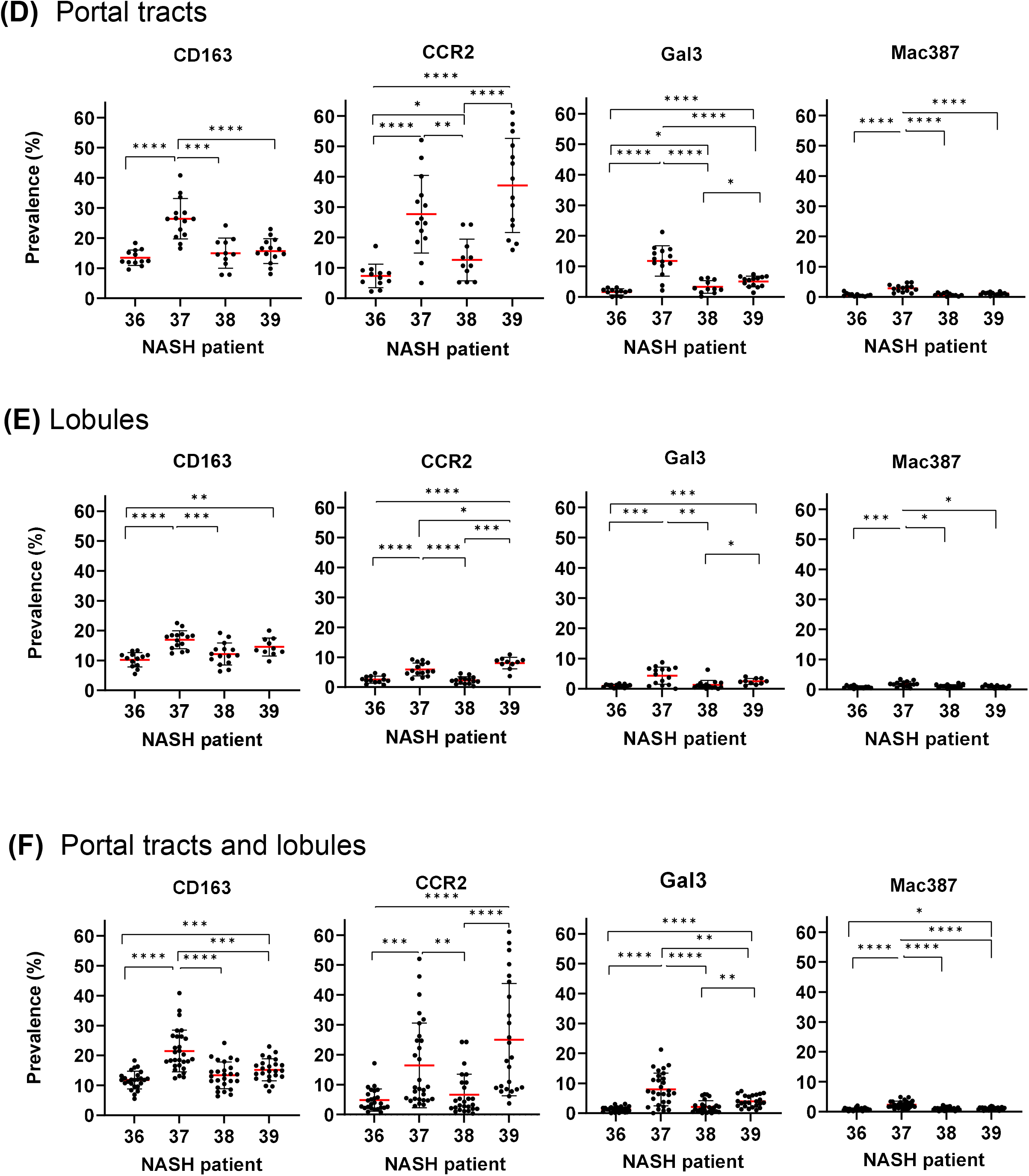
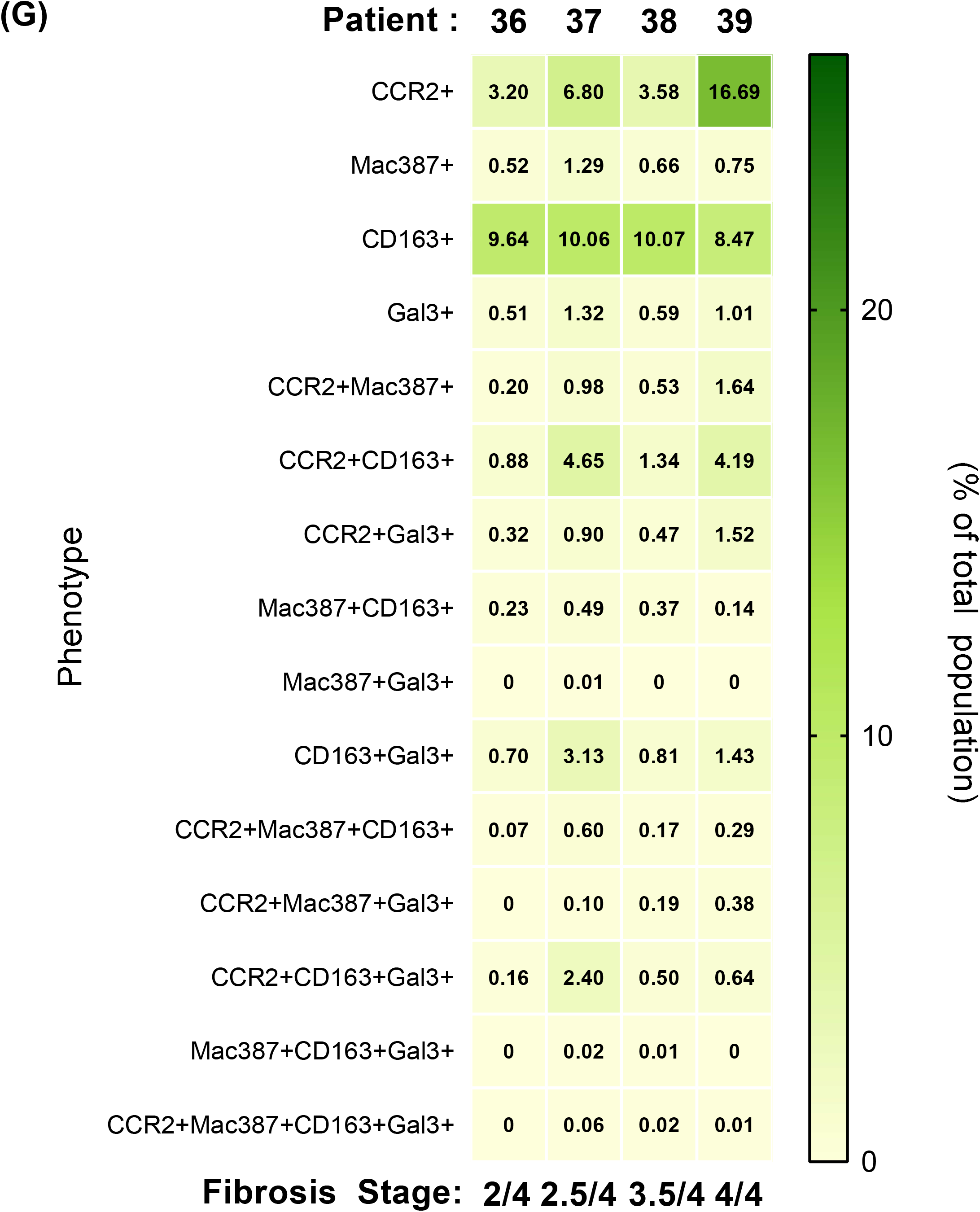
Quantitative comparison of four different patients with NASH showed heterogeneity in intrahepatic macrophages and variable expression of therapeutic targets. Images acquired from individual patients (patients 36-39 as shown in Figure 7) were analyzed using custom-designed Visiopharm applications to calculate the percentage of marker-positive cells after normalization by the total number of cells detected (DAPI+). **(A-C)**: The heat map shows the mean percent prevalence of marker-positive cells in the patients with different fibrosis stages in regions enriched with **(A)** portal tracts, **(B)** lobules, **(C)** portal tracts and lobules combined. **(D-F)**: Next, we used GraphPad Prism to compare the statistical differences between the prevalence of each marker among the patients with different fibrosis stages. Patients 37 and patient 39, who had active HCV and NASH, had the highest prevalence of Galectin-3 and CCR2 expression compared to the other patients. All data are presented as means (red line) with significance calculated using unpaired t-test (*p ≤ 0.05; ** p ≤ 0.01; ***p ≤ 0.001; ****p≤ 0.0001). Each black dot (•) represents the prevalence of a particular maker in an individual image. **(G)**: The percentages of the different phenotypes resulting from the therapeutic panel was also calculated for the portal tracts and lobules. CCR2+CD163+ and CD163+Gal3+ phenotypes were increased in patients 37 and 39 relative to the value of the other patients.

## DISCUSSION

NASH can lead to cirrhosis, end-stage liver disease, and HCC and it is now the number one cause of liver transplantation in women and the second most common cause in men [1]. Not only is NASH increasing in prevalence, but it is also often clinically silent, and when diagnosed, is challenging to treat [33, 34]. Several antifibrotic therapies in clinical trials target intrahepatic macrophages to treat NASH (**Table S1**) without any knowledge of the patient’s underlying hepatic microenvironment. Initial trials for some of the most promising agents, such as cenicriviroc, terminated early due to lack of efficacy. Initial results of the CENTAUR trial showed that ∼ 20% of patients had improvement of fibrosis (≥ 1 stage) without affecting steatosis or inflammatory activity associated with NASH at the end of year 1. At the end of year 2, no further improvement in fibrosis was observed [7]. Since cenicriviroc inhibits infiltration of pro-fibrotic macrophages into the liver via antagonism of CCR2/CCR5 and has been reported to decrease inflammation, steatosis, and fibrosis [7, 11, 12, 30, 35, 36], these results raised several questions. Why did patients with advanced fibrosis have a more pronounced treatment response? Did all patients with biopsy-proven NASH have infiltrating macrophages in their livers? Why did such a small percentage of patients respond to therapy? Even though serial liver biopsies were collected from these patients at baseline, the end of year 1, and the end of year 2, these were only used to compare histologic changes (i.e., NAS scores) and fibrosis stages [7]. We developed this study to provide clues to the questions above and to improve understanding of these complex cells within the human hepatic microenvironment. These results also provide strong evidence that we may be able to predict who will respond to treatment based on the phenotypes and level of marker expression present in the liver.

First, we compared a group of well-matched patients with biopsy-confirmed NASH (**Table S3**) and extracted RNA from unstained FFPE slides for nCounter analysis (**Fig 2**). Gadd et al. previously reported that CD68+ macrophages were increased in the portal tracts of patients with NAFLD and were one of the earliest changes detected [31] and another study using digital spatial profiling revealed greater mRNA expression of CD68 in patients with advanced fibrosis due to NASH [37]. Interestingly, we did not detect significant increases in the main macrophage-related genes (e.g., CD68 and CD163) as we expected. However, several relevant genes related to this study were upregulated and significantly increased in the patients with advanced fibrosis including CCL2, CCL5, CCR2, and Galectin-3 (LGALS3). Even though we showed variability in the protein expression of several of these markers *in situ* in the hepatic microenvironment (**Figures 7-8**), in a larger group of patients, overall higher expression was observed in patients with advanced fibrosis due to NASH. However, homogenization of the liver tissue was unable to detect important differences in individual patients, phenotype percentages in the context of the hepatic architecture, or significant spatial relationships. Due to the above previous studies and initial nanoString results, additional experiments were conducted using spectral imaging microscopy; an approach that can analyze macrophage populations in situ with preservation of hepatic architecture.

Since cenicriviroc resulted in a more pronounced response in patients with advanced fibrosis [7], we hypothesized that patients in this study with bridging fibrosis/cirrhosis would have increased prevalence of infiltrating macrophages, enhanced spatial relationships, enrichment of macrophage phenotypes, and more expression of treatment targets, like CCR2.

In addition, since only a small percentage of patients showed improvement after cenicriviroc treatment, we also predicted that there would be variability in the prevalence of certain phenotypes and marker expression between individual patients with NASH. As expected, multispectral images and phenotype profiles acquired from the patients with advanced fibrosis showed increased numbers of macrophages within the portal tracts and lobules (**Figure 3**). Liver biopsies from patients with advanced fibrosis also contained differential macrophage distribution and phenotypes as highlighted in the t-SNE and phenotype profile plots (**Figures 3C, 3F, and 3G**). Patients with advanced fibrosis had a higher prevalence of CD68+, CD163+, and Mac387+macrophages, while several phenotypes, such as CD14+CD16+, and CD14+CD16+CD163+, showed similar prevalence in both groups of patients. However, differences when comparing the groups of patients were often subtle which we determined was due to heterogeneity in the individual patients within each group. CD68+ and Mac387+ phenotypes have also been shown to be increased in patients with chronic liver disease when compared to controls [11]. Although we expected to see some variability in the prevalence of Mac387 expression in the individual patients, we did not expect the two patients with minimal fibrosis (patients 27 and 29) to have a higher prevalence of infiltrating macrophages in the liver than most of the patients with advanced fibrosis (see **Figure 4A)**. These two patients also later developed features of portal hypertension, including elevated prothrombin time, thrombocytopenia, and splenomegaly, approximately 7.6 and 9.4 years later (see **Table S6**).

Patients 33 and 34 in the advanced fibrosis group also had a higher prevalence of Mac387 expression when compared to the other patients (**Figure 4B**). Patient 32 was reported deceased due to end-stage liver disease at follow-up 3.2 years later, and her baseline liver biopsy did not show an increased prevalence of the macrophage phenotypes evaluated (see **Figure 4B**). Patient 34 had a high prevalence of numerous phenotypes within in the liver, including CD16+, CD163+, Mac387+, CD16+CD163+, CD68+CD16+CD163+, and CD14+CD16+CD163+, and was reported to be deceased from a bowel rupture due to underlying ischemic colitis, 4.9 years after her baseline biopsy.

Several other known profibrotic phenotypes showed higher percentages in patients with advanced fibrosis, including CD16+, CD163+, CD16+CD163+, and CD68+CD16+CD163+ populations (**Figures 3 and 4**) [15]. Human CD16+ macrophages are an anti-inflammatory phenotype that has patrolling activities on the vascular endothelium and transmigrates in a vascular adhesion protein-1-(VAP-1) and CX3CR1-dependent manner (thought to be independent of CCR2), allowing localization at sites of inflammation and fibrosis [14, 38, 39]. Several phenotypes that expressed CD16 alone or in combination with other markers were present in the patients with advanced fibrosis and this phenotype may be another worth exploring for targeted therapies. Another well-recognized human intermediate inflammatory monocyte/macrophage present in both groups of patients with NASH is the CD14+CD16+ subset. This population accumulates in other models of chronic liver diseases and is derived from the recruitment of blood monocytes or after local differentiation of classical CD14++(bright)/CD16-(dim) monocytes in response to TGFβ and IL10 that are present in the chronically inflamed liver [15]. This intermediate monocyte-derived phenotype has features of both macrophages and dendritic cells, has phagocytic functions, can act as antigen-presenting cells, and secretes cytokines (e.g., IL6, IL8, IL1β, IL13) and chemokines (e.g., CCL1/2/3/5) that modulate inflammation and fibrogenesis [15]. Higher CD14+ expression (without other markers) was observed in the patients with minimal fibrosis. CD14 has been shown to be present in more tolerogenic livers and can be found on type 2 liver sinusoidal endothelial cells [22, 40, 41]. Higher expression of CD14 lining the sinusoids was observed in multispectral images from the patients with minimal fibrosis, while lower expression was observed in patients with advanced fibrosis (see **Figure 3**). However, there was marked variability in the expression of CD14 in the individual patients.

The differences in macrophage phenotypes and prevalence showed that quantities of cells varied within the hepatic microenvironment between groups and individual patients. However, these initial results did not provide any information about the spatial relationships of these phenotypes within the liver. Since spatial analysis of the tumor microenvironment has predicted drug efficacy and clinical outcome in patients with HCC and cholangiocarcinoma, respectively [42, 43], we applied similar approaches to patients with NASH. We used UMAPs, G-function, and Giotto enrichment/depletion spatial analyses to quantify the mixing and/or infiltration of macrophage phenotypes within the liver. The interaction of Mac387+ and CD68+ cells, either with each other or with other markers, was significantly increased in the patients with advanced fibrosis (see **Figure 5**). Huang et al. also showed that these two phenotypes increase in the liver in patients with chronic liver disease [11] as we also showed in this study. It seems obvious that cells that are increased in number in a tissue site will have more opportunities to be near one another. However, even though we evaluated the spatial relationships of all 30 phenotypes identified in this study, many of which had increased percentages in patients with advanced fibrosis, only the interaction between Mac387+ and CD68+ macrophages reached statistical significance (see **Figure 5**). Analysis of individual patients in the minimal fibrosis group showed that only two patients had increased enrichment of Mac387+ macrophages (**Figures 6A and 6C**, patients 27 and 29), and two of these patients later developed features of portal hypertension (see **Figure 6** and **Table S6**). Three patients (**Figure 6B**, patients 31, 32, and 34) within the advanced fibrosis group showed more enrichment interactions than the other patients. One of these patients later developed portal hypertension (patient 31), one died of ESLD (patient 32), and one was deceased due to a bowel rupture (patient 34); all within a few years of their baseline biopsies, (see **Figure 6B** and **Table S6**). Larger studies will need to be conducted to confirm that spatial relationships within the hepatic microenvironment are able to predict clinical outcomes in medical liver disease. However, these interactions deserve further investigation since Mac387+ cells are known to mediate hepatic fibrosis and are one of the main targets of drugs like cenicriviroc.

The amount of heterogeneity among individual patients with NASH, particularly with phenotypes that are known to be pro-fibrotic (e.g., Mac387+ and CD163+), prompted us to evaluate the next four patients diagnosed with NASH by liver biopsy at our institution. We conducted spectral imaging analysis in addition to routine NAS grading and fibrosis staging. A single unstained liver biopsy slide from each patient was stained with a therapy-related multiplex panel that included antibodies against CD163, Galectin-3, CCR2, and Mac387 (see **Figure 7-8**). Multiplex images from these patients showed obvious differences within both the portal tracts and lobules that were not always dependent on the fibrosis stage. Of these four patients with varied stages of fibrosis, patient 37, a patient with early bridging fibrosis (stage 3/4) had the highest prevalence of CD163+ macrophages and expression of Galectin-3. CD163+ was expressed at higher levels in both the lobules and portal tracts when compared to Mac387+ macrophages (see **Figure 8A-F**). Patient 39, a patient that had active hepatitis C and NASH, had the highest expression of CCR2. Since these were recent biopsies, outcomes in these patients are yet to be determined. However, it is known that patients with increased PD-L1+ host cells within the hepatic microenvironment are more likely to respond to immunotherapy against this target, and a recent study by Lu et al. in *Gut* reported that selective modulation of macrophages restores antitumoral properties in patients with HCC [44]. If macrophages can be “selectively modified” in the livers of patients that already have cancer, they most certainly can be modified earlier in the course of the disease where poor outcomes like cirrhosis and HCC have a chance of being prevented.

A major strength of this study is that we analyzed the human liver microenvironment *in situ* in biopsies from real patients with NASH. So many studies of the hepatic microenvironment and immune response have been conducted *in vitro* and in mouse models, including several that led to cenicriviroc therapy for NASH [45]. It is well known that mice differ a lot from humans [46] [47]. An elegant study by Jiang et al. completed a comparative study of the mouse, human, and humanized mouse livers in fatty liver disease, and showed that gene expression in mice and humans is very different, with only 1524 genes commonly regulated [47]. In addition, it takes years to develop cirrhosis and HCC when you have chronic liver disease, so replicating the duration of the disease is not feasible when using animal models.

Studies in humans, although more translational and often more clinically relevant, also pose challenges. First, mechanistic studies are nearly impossible in humans, whereas they are common in mouse models. Second, as we observed in these patients, macrophages within the hepatic microenvironment vary substantially and each patient has their own unique population. The small sample size is an obvious limitation of this study. However, this initial study was used to develop effective imaging analysis tools to characterize intrahepatic macrophages *in situ* in human medical liver biopsies. These data also enabled us to customize Visiopharm algorithms to optimized multiplex panels, enabling higher throughput analysis of more patients and other chronic liver diseases in the future. Another limitation of this study is that liver biopsy tissue is required. Liver biopsies are still considered the gold standard for diagnosis and fibrosis staging in patients with NAFLD/NASH according to AASLD guidelines [4]. Since other less invasive tools such as Fibroscan and tissue elastography are being more frequently utilized [48], biopsies are collected less frequently, which makes the development of precision medicine approaches even more challenging. Patients with cancer routinely provide tissue samples for molecular testing and to determine the expression of targets by immunohistochemical staining prior to treatment with chemo- and immunotherapies [49, 50]. Even though liquid biopsy is being used successfully for cancers, there has been limited success with systemic biomarkers for inflammatory and infectious diseases [51, 52]. We need to fully understand the complexity of the hepatic microenvironment before reliable clinical surrogates can be used for targeted therapies. If a small piece of liver tissue could predict clinical outcomes (before they happen), predict treatment response while minimizing side-effects, and detect changes in hepatic microenvironments that are abnormally pro-fibrotic or pro-cancerous, the benefits of liver biopsy may outweigh the risks.

In summary, if we can learn how to modulate the hepatic microenvironment earlier in the course of the disease before cirrhosis or HCC develop, we may be able to prevent bad outcomes before it is too late to intervene. We hope the results of this study will inspire the development of more personalized therapeutic approaches for NASH and look forward to the future of translational medicine to treat medical liver diseases.

## Data Availability

Data analyzed during the current study are available upon reasonable request to the authors.

## ACKNOWLEDGEMENTS

Sam Diaz de Leon and Blanca Hernandez provided secretarial support and assistance with obtaining the archived liver biopsy tissue blocks. We thank Drs. John Vierling and Frank Tacke for their expert feedback regarding figures in this paper. Shana White, clinical study coordinator, assisted with chart review, IRB approval, and demographic information.

## FIGURE LEGENDS

**Supplementary Figure 1. Representative liver biopsies collected from patients with either minimal or advanced fibrosis due to NASH and collagen proportionate area (CPA) calculation. (A-B):** Masson’s trichome stains were used to stage the fibrosis in patients with NASH. A group of patients with minimal fibrosis **(A)** (fibrosis stage: 0-1 out of 4; n = 6, patients 25-30) were compared to a group with advanced fibrosis **(B)** in representative patients with NASH (fibrosis stage: 3.5 or 4 out of 4; n = 5, patients 31-35) [23]. **(C)**: We developed an algorithm using Visiopharm (imaging analysis software) to quantify collagen (CPA) more objectively in the liver biopsies. CPA areas were determined by detecting the area of the Masson’s trichrome stains divided by the total surface area of the biopsy to calculate the percentage of fibrosis. The percent CPA = ∑fibrotic area/∑total area x 100. Patients with advanced fibrosis had significantly (p<0.004) increased CPA when compared to patients with minimal fibrosis. The Mann-Whitney test was used to compare differences between the groups. *p < 0.05; **p < 0.01; ***p < 0.001; ****p < 0.0001. **Table S5** shows all the histopathologic findings in the patient biopsies (n = 11), including fibrosis stages using established criteria for grading/staging liver biopsies for NASH [23].

## List of Abbreviations

AIH: autoimmune hepatitis
ALT: alanine transaminase
ALP: alkaline phosphatase
CAP: College of American Pathologists
CPA: collagen proportionate area
ESLD: end stage liver disease
ESRD: end stage renal disease
FFPE: formalin-fixed, paraffin-embedded
H&E: hematoxylin and eosin
HCC: hepatocellular carcinoma
HCV: hepatitis C virus
IHC: immunohistochemical
NAFLD: non-alcoholic fatty liver disease
NAS: NAFLD activity scores
NASH: nonalcoholic steatohepatitis
ROI: region of interest
TSA: tyramide signal amplification
t-SNE: t-distributed stochastic neighbor embedding
MDACC: MD Anderson Cancer Center

