## Supplementary data for "Patients with fibrosis from non-alcoholic steatohepatitis have heterogeneous intrahepatic macrophages and therapeutic targets"

**(Table S1).** Promising antifibrotic therapies that target macrophages in patients with non-alcoholic steatohepatitis

| Drug | Target | Mechanism of Action in the Liver | Clinical Trial (https://Clinicaltrials.gov) |
|---|---|---|---|
| **Cenicriviroc (CVC)** | CCR2/CCR5 | Dual CCR2/CCR5 antagonist; on monocytes, Kupffer cells and HSCs inhibits systemic Mac infiltration | CENTAUR (NCT02217475): improvement in fibrosis score across year 1 and 2 (PMID: 31943293)<br>AURORA (NCT03028740): terminated early due to lack of efficacy based on results of planned interim analysis |
| **Cenicriviroc (CVC) + Tropifexor** | CCR2/CCR5 + Fibroblast Growth Factor (FGF)19 | Dual CCR2/CCR5 antagonist; on monocytes, Kupffer cells and HSCs inhibits systemic Mac infiltration; FGF19 analog | TANDEM (NCT03517540): phase II completed; sustained improvement in ALT and bodyweight, but no treatment benefit of combination therapy greater than monotherapy |
| **Galactoarabino-rhamnogalaturonan (GR-MD-02)** | Galectin 3 | Galectin 3 antagonist; inhibits HSC and Kupffer cell activation | GR-MD-02 (NCT02421094): phase II completed, moving to phase III (PMID: 27778367) |
| **GB1211** | Galectin 3 | Galectin 3 antagonist; inhibits HSC and Kupffer cell activation | GULLIVER-2 (NCT05009680): active phase II trial |
| **Obeticholic acid (OCA)** | Farnesoid X receptor (FXR)/NR1H4 | FXR agonist; on Macs and HSCs, expressed in the liver and gut (27) | REGENERATE (NCT02548351): active phase III (PMID: 34274514)<br>REVERSE (NCT03439254): active phase III |
| **Resmetirom** | Thyroid Hormone Receptor β (THR-β) | THR-β Agonist; expressed on cholangiocytes, macrophages, and some hepatocytes (PMID: 24215914) | MAESTRO-NASH (NCT03900429 and NCT05500222): active phase IIIa and b studies<br>MAESTRO-NAFLD 1 (NCT04951219): active phase III |
| **Aramchol (3β-Arachidyl amido cholanoic acid)** | (Stearoyl Co-A Desaturase 1 or SCD-1) | SCD-1 Partial Inhibitor; expressed on HSCs, macrophages, and hepatocytes (PMID: 23747827) | Aramchol003 (NCT01094158): completed phase II study (PMID: 24815326)<br>ARRIVE (NCT02684591): completed phase II for HIV-associated NAFLD; failed to meet primary/secondary endpoints (PMID: 31013363)<br>ARREST (NCT02279524): phase IIb for NASH showed improvement in fibrosis stage1 without worsening of NASH and improved ALT (PMID: 34621052)<br>ARMOR (NCT04104321): suspended phase III early, due to promising results with histological improvement in 50% of participants |
| **Denifanstat (TVB 2640)** | Fatty acid synthase (FASN) | FASN inhibitor; expressed in hepatocytes, macrophages, and HCC cells (PMID: 29463677) | FASCINATE-1 (NCT03938246): completed phase II study (no results posted) (PMID: 34310978)<br>FASCINATE-2 (NCT04906421): active phase IIb study |

**(Table S2).** Antibodies and optimized multiplex conditions used to identify human macrophage phenotypes and drug targets in human formalin-fixed, paraffin-embedded liver biopsy tissues

| Antibody | Vendor/Clone/Isotype | Dilution | Incubation Time (min) | Antigen Retrieval Buffer | Opal/Dilution | Reaction Position |
|---|---|---|---|---|---|---|
| | | | **Macrophage phenotype Panel (manual)** | | | |
| **CD68** | Bio*/KP1/mouse-IgG1κ | RTU** | 60 | AR9 (Ak***) | 520/1:300 | 1 |
| **MAC387** | Dako/Mac387/mouse-IgG1κ | 1:200 | 30 | AR6 (Bio*) | 690/1:100 | 2 |
| **CD16** | Abcam/EPR16784/rabbit-IgG | 1:500 | 60 | AR6 (Bio*) | 620/1:600 | 3 |
| **CD14** | Abcam/EPR3653/rabbit-IgG | 1:500 | 45 | AR6 (Bio*) | 540/1:600 | 4 |
| **CD163** | Leica/10D6/mouse-IgG1 | RTU** | 30 | AR6 (Bio*) | 650/1:100 | 5 |
| **DAPI** | Ak*** | 15uL/mL | 4 | AR6 (Bio*) | - | 6 |
| | | | **Macrophage therapy related target panel (automated)** | | | |
| **CCR2** | Abcam/7A7/mouse-IgG | 1:200 | 20 | CC1 | 650/1:150 | 1 |
| **MAC387** | Dako/Mac387/mouse-IgG1κ | 1:500 | 20 | CC2 | 690/1:100 | 2 |
| **Galectin3** | Millipore Sigma/9C4/mouse-IgG1 | RTU | 24 | CC1 | 570/1:100 | 3 |
| **CD163** | Cell Marque/MRQ-26/mouse-IgG | RTU | 32 | CC2 | 520/1:100 | 4 |
| **DAPI** | Roche | RTU | 16 | - | - | 5 |

Bio*, Biogenex; ** RTU, ready to use; Ak***, Akoya Biosciences

**(Table S3).** Demographic and laboratory data for matched nonalcoholic steatohepatitis patients with minimal and advanced fibrosis analyzed with nCounter

| | Patients with minimal fibrosis | | | | | | | | | | | | Patients with advanced fibrosis | | | | | | | | | | | | |
|---|---|---|---|---|---|---|---|---|---|---|---|---|---|---|---|---|---|---|---|---|---|---|---|---|---|
| **Patient #** | **1** | **2** | **3** | **4** | **5** | **6** | **7** | **8** | **9** | **10** | **11** | **12** | **13** | **14** | **15** | **16** | **17** | **18** | **19** | **20** | **21** | **22** | **23** | **24** | **p-value** |
| **Age range** | 30s - 60s | | | | | | | | | | | | 30s - 60s | | | | | | | | | | | | ns |
| **Sex** | F | F | F | M | M | F | F | F | M | F | M | M | F | F | F | M | M | F | F | F | M | F | M | M | - |
| **Race** | W | W | W | W | W | H | H | H | W | W | W | H | W | W | H | W | H | H | H | H | W | H | H | W | - |
| **ALT** | 37 | 40 | 35 | 92 | 85 | 35 | 83 | 52 | 155 | 38 | 42 | 68 | 43 | 54 | 50 | 58 | 99 | 39 | 167 | 22 | 118 | 81 | 64 | 61 | ns |
| **AST** | 18 | 31 | 25 | 55 | 48 | 35 | 53 | 29 | 79 | 36 | 24 | 43 | 49 | 84 | 41 | 50 | 74 | 65 | 149 | 16 | 65 | 69 | 101 | 77 | ** |
| **ALP** | 97 | 59 | ND | 64 | 77 | 135 | 104 | 92 | 82 | 97 | 75 | 75 | 119 | 164 | 150 | 42 | 81 | 178 | 194 | 102 | 88 | 90 | 104 | 80 | ns |
| **Bilirubin** | 0.1 | 0.4 | 0.3 | 0.5 | 0.3 | 0.1 | 0.6 | 0.3 | 1.1 | 0.1 | 0.4 | 0.4 | 0.5 | 0.7 | 0.6 | 1.4 | 1.2 | 1.3 | 1.5 | 0.5 | 0.8 | 0.5 | 4 | 0.4 | ns |
| **Albumin** | 4.1 | 3.6 | 4.1 | 4.7 | 4.5 | 3.9 | 3.7 | 4.6 | 4.6 | 4.6 | 4.2 | 4.4 | 3.7 | 4.5 | 4.1 | 3.5 | 4.1 | 3.3 | 3.5 | 4.2 | 5 | 3.6 | 3.1 | 4.2 | ns |
| **Platelets** | 275 | 314 | 258 | 221 | 262 | 264 | 248 | 310 | 192 | 300 | 264 | 273 | 158 | 82 | 207 | 121 | 133 | 72 | 75 | 257 | 206 | 183 | 73 | 292 | **** |
| **BMI** | 49.5 | 41.6 | 43.8 | 36.5 | 46.8 | 37.5 | 50.4 | 29 | 31.86 | 38.05 | 35.5 | 32.79 | 34.6 | 44.39 | 36.73 | 40.8 | 46.52 | 30 | 39.92 | 39 | 39.5 | 45 | 36.02 | 50.22 | ns |
| **NAS score** | 3 | 3 | 3 | 5 | 4 | 4 | 2 | 1 | 3 | 2 | 5 | 4 | 3 | 4 | 5 | 3 | 5 | 3 | 5 | 6 | 6 | 7 | 4 | 7 | * |
| **Fibrosis stage** | 1 | 1 | 1 | 1 | 1 | 1 | 1 | 1 | 1 | 1 | 1 | 1 | 4 | 4 | 4 | 4 | 4 | 4 | 4 | 4 | 4 | 4 | 4 | 4 | **** |

Normal range: ALT, 9-51 U/L; AST,13-40 U/L; ALP, 34-122 U/L; Bilirubin, 0.1-1.1 mg/dL; Albumin, 3.5-5 g/dL; Platelets,166-358 x1000/uL.
Abbreviations: F, female; M, male; BMI, body mass index; W, White; H, Hispanic; NAS, NAFLD activity score (0-8); ns, not significant. An unpaired t-test was used to compare differences between the groups. *p < 0.05; **p < 0.01; ***p < 0.001; ****p < 0.0001.

(**Table S4**). Gene expression in matched nonalcoholic steatohepatitis patients with minimal and advanced fibrosis analyzed with nCounter.

| Gene | Fold Change | P-value | Gene | Fold Change | P-value |
|---|---|---|---|---|---|
| **"M1" Classical Activation** | | | **Chemokines** | | |
| CD68 | 1.092 | 0.660 | CCR2 | 2.006 | 0.003 |
| CD86 | 0.951 | 0.767 | CCR5 | 1.323 | 0.108 |
| CD80 | 1.101 | 0.599 | CXCL5 | 1.368 | 0.257 |
| IL1R1 | 1.069 | 0.604 | CCL18 | 2.393 | 0.008 |
| TLR2 | 0.708 | 0.216 | **Cytokines** | | |
| TLR3 | 1.162 | 0.211 | IL1B | 0.976 | 0.899 |
| TLR4 | 0.861 | 0.234 | TNF | 1.318 | 0.134 |
| TLR7 | 0.885 | 0.448 | IL12A | 0.788 | 0.406 |
| TLR9 | 1.143 | 0.615 | IL23A | 0.884 | 0.664 |
| FCGR1A/CD64 | 1.198 | 0.336 | IFNG | 0.924 | 0.756 |
| CD14 | 0.811 | 0.280 | IL18 | 1.273 | 0.174 |
| MARCO | 0.634 | 0.051 | IL10 | 0.701 | 0.216 |
| HLA-DRA | 2.188 | 0.022 | **Signaling Molecules** | | |
| HLA-DRB3 | 1.369 | 0.037 | Btk | 1.153 | 0.410 |
| HLA-DRB4 | 3.873 | 0.213 | SOCS1 | 1.145 | 0.480 |
| **"M2" Alternative Activation** | | | MyD88 | 0.804 | 0.114 |
| CD163 | 0.656 | 0.003 | TICAM1 | 1.146 | 0.211 |
| CD206/MRC1 | 0.890 | 0.496 | AKT3 | 1.295 | 0.014 |
| FCER2/CD23 | 1.185 | 0.426 | SYK | 1.840 | 0.001 |
| CD209/DCSIGN | 0.976 | 0.877 | PIK3CD | 1.894 | 0.001 |
| CLEC7A | 1.402 | 0.002 | PIK3CG | 2.025 | <0.01 |
| MSR1/CD204 | 1.234 | 0.141 | HRAS | 1.117 | 0.428 |
| TLR1 | 1.628 | 0.001 | MAPK1 | 1.501 | 0.058 |
| TLR8 | 1.198 | 0.264 | **Fibrosis** | | |
| IL4R | 1.041 | 0.721 | STAT3 | 0.790 | 0.180 |
| FCER1G | 0.988 | 0.953 | TGFB1 | 1.712 | 0.004 |
| IL10RA | 1.638 | 0.009 | TGFB2 | 1.313 | 0.160 |
| CSF1R | 0.853 | 0.371 | TREM1 | 0.931 | 0.817 |
| PDCD1LG2/PDL2 | 1.118 | 0.469 | MERTK | 0.842 | 0.326 |
| CD274/PDL1 | 0.820 | 0.239 | LGALS3 | 3.658 | <0.01 |
| CD36 | 0.908 | 0.464 | **Transcription Factors** | | |
| CD1B | 1.131 | 0.613 | STAT1 | 1.750 | 0.057 |
| FCGR3A/CD16 | 1.023 | 0.903 | IRF1 | 1.446 | 0.013 |
| **Chemokines** | | | IRF5 | 1.130 | 0.586 |
| CCL2 | 2.423 | 0.005 | IRF8 | 1.343 | 0.059 |
| CCL3 | 0.950 | 0.784 | STAT5B | 1.171 | 0.190 |
| CCL4 | 1.311 | 0.334 | STAT6 | 1.247 | 0.058 |
| CCL5 | 1.786 | 0.007 | IRF4 | 2.298 | <0.01 |
| CCL8 | 1.066 | 0.778 | NFKB1 | 1.147 | 0.297 |
| CCL11 | 0.756 | 0.285 | PPARG | 1.013 | 0.967 |
| CXCL10 | 2.250 | 0.090 | MAF | 0.840 | 0.214 |
| CXCL9 | 4.409 | 0.237 | **Enzymes** | | |
| CCL17 | 0.988 | 0.946 | IDO1 | 1.505 | 0.034 |
| CCL22 | 1.117 | 0.647 | PTGS2 | 1.263 | 0.394 |
| CCL24 | 0.761 | 0.311 | ARG1 | 0.581 | 0.002 |

Note: Gene expression analysis was performed in patients with minimal (n = 12; fibrosis stage = 1/4) and advanced fibrosis (n = 12; fibrosis stage = 4/4) using the PanCancer immune profiling panel, the nCounter sprint profiler and the nSolver advanced analysis module. Gene names and values in green (upregulated) and red (downregulated) in the advanced vs minimal fibrosis group. $p < 0.05$ was considered significant.

**(Table S5).** Demographic and laboratory data for nonalcoholic steatohepatitis patients at the time of baseline biopsy

| | Patients with minimal fibrosis | | | | | | Patients with advanced fibrosis | | | | | |
|---|---|---|---|---|---|---|---|---|---|---|---|---|
| **Patient #** | **25** | **26** | **27** | **28** | **29** | **30** | **31** | **32** | **33** | **34** | **35** | **p-value** |
| **Age range** | 20s - 50s | | | | | | 40s - 70s | | | | | ns |
| **Sex** | F | F | F | M | F | F | M | F | M | F | F | - |
| **ALT** | 88 | 155 | 38 | 68 | 40 | 30 | 99 | 13 | 34 | 20 | 58 | ns |
| **AST** | 44 | 79 | 36 | 40 | 31 | 28 | 74 | 49 | 31 | 34 | 67 | ns |
| **ALP** | 118 | 82 | 97 | 93 | 59 | 127 | 81 | 146 | 58 | 78 | 80 | ns |
| **Bilirubin** | 0.2 | 1.0 | 0.1 | 0.6 | 0.4 | 0.3 | 1.1 | 4.6 | 0.5 | 0.9 | 0.9 | ** |
| **Albumin** | 4.6 | 4.3 | 4.6 | 4.5 | 3.6 | 4.3 | 4.1 | 3.9 | 4.2 | 4.0 | 4.0 | ns |
| **Platelets** | 396 | 192 | 268 | 264 | 314 | 187 | 130 | 149 | 292 | 210 | 173 | ns |
| **BMI** | 35 | 33 | 35 | 35 | 35 | 36 | 47 | 31 | 45 | 32 | 32 | ns |
| **NAS (scores)** | 4 (70-3-1-0) | 2 (60-2-1-0) | 2 (10-1-1-0) | 5 (40-2-2-1) | 4 (35-2-1-1) | 3.5 (20-1-1.5-1) | 4 (35-2-1-1) | 5 (60-2-2-1) | 6 (10-1-3-2) | 3 (<5-0-2-1) | 5 (20-1-2-2) | ns |
| **Fibrosis stage** | 1 | 1 | 0-1 | 0-1 | 0-1 | 1 | 3-4 | 4 | 4 | 4 | 4 | *** |
| **CPA** | 0.94 | 6.06 | 0.53 | 0.35 | 0.19 | 1.68 | 20.12 | 6.94 | 25.87 | 25.56 | 13.11 | *** |

Normal range: ALT, 9-51 U/L; AST,13-40 U/L; ALP, 34-122 U/L; Bilirubin, 0.1-1.1 mg/dL; Albumin, 3.5-5 g/dL; Platelets,166-358 x1000/uL.
Abbreviations: F, female; M, male; BMI, body mass index; NAS, NAFLD activity score (0-8); CPA, collagen proportionate area; ns, not significant. An unpaired t-test was used to compare differences between the groups. *p < 0.05; **p < 0.01; ***p < 0.001; ****p < 0.0001.

### Figure S1

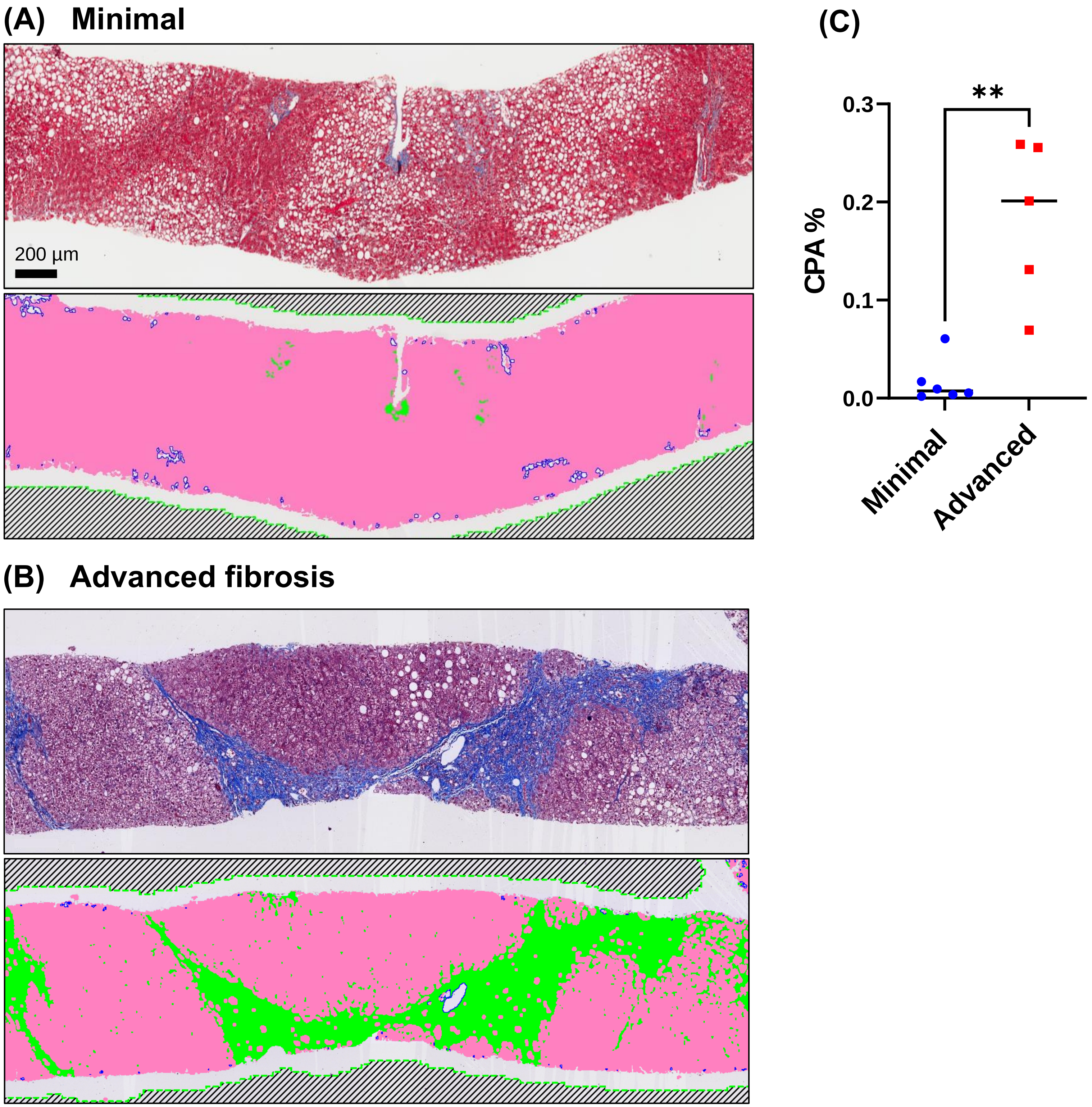

(A) Minimal
200 µm
(B) Advanced fibrosis
(C)
**
CPA %
0.3
0.2
0.1
0.0
Minimal
Advanced

**(Table S6).** Clinical outcome for patients with nonalcoholic steatohepatitis at the time of baseline biopsy

| | **Patients with minimal fibrosis** | | | | | | **Patients with advanced fibrosis** | | | | |
|---|---|---|---|---|---|---|---|---|---|---|---|
| **Patient #** | **25** | **26** | **27** | **28** | **29** | **30** | **31** | **32** | **33** | **34** | **35** |
| **Age at Baseline Biopsy** | 20s - 50s | | | | | | 40s - 60s | | | | |
| **Year of Baseline Biopsy** | 2009 - 2014 | | | | | | 2006 - 2017 | | | | |
| **Duration of F/U after Baseline Biopsy (years)** | 13 | 2 | 12 | 6 | 13 | 13 | 5 | 3 | 12 | 4 | 16 |
| **Portal Hypertension** | No | No | **Yes** | No | **Yes** | No | **Yes** | **Yes** | No | No | No |
| **Cancer Diagnosis** | No | No | No | No | No | No | No | No | No | No | No |
| **Transplanted** | No | No | No | No | No | No | No | Waitlisted | No | No | No |
| **Deceased** | No | No | No | No | No | No | No | **Yes**, 2011 | No | **Yes**, 2013 | No |
| **Cause of Death** | n/a | n/a | n/a | n/a | n/a | n/a | n/a | **ESLD** | n/a | Bowel rupture | n/a |
| **Days Between Baseline Biopsies to Portal HTN/CA/Death** | n/a | n/a | 2,775 | n/a | 3,431 | n/a | 0 | **1,204** | n/a | 1,533 | n/a |

Abbreviations: F/U, follow up; HTN, hypertension; CA, cancer; n/a, not applicable

**(Table S7).** Demographic and laboratory data for additional nonalcoholic steatohepatitis patients at the time of baseline biopsy

| | Patients with variable fibrosis stages | | | |
|---|---|---|---|---|
| **Patient #** | **36** | **37** | **38** | **39** |
| **Age decile** | 50 - 59 | 70 - 79 | 60 - 69 | 70 - 79 |
| **Sex** | M | F | F | F |
| **Race** | H | H | W | W |
| **ALT** | 38 | 191 | 28 | 47 |
| **AST** | 30 | 189 | 41 | 57 |
| **ALP** | 61 | 136 | 66 | 90 |
| **Bilirubin (uMol/L)** | 0.7 | 0.5 | 0.7 | 0.6 |
| **Albumin (g/L)** | 4.8 | 4.3 | 3.0 | 4.4 |
| **Platelet ct (x1000)** | 295 | 239 | 66 | 202 |
| **BMI** | 40.75 | 33 | 31 | 26 |
| **Percent steatosis** | 50 | 30 | 30 | 40 |
| **NAS (scores)** | 3<br>(1-1-1) | 6<br>(1-3-2) | 4<br>(2-1-1) | 4<br>(2-2-0) |
| **Fibrosis stage** | 2 | 3 | 4 | 4 |

Normal range: ALT, 9-51 U/L; AST,13-40 U/L; ALP, 34-122 U/L; Bilirubin, 0.1-1.1 mg/dL; Albumin, 3.5-5 g/dL; Platelets,166-358 x1000/uL.
Abbreviations: F, female; M, male; W, White; H, Hispanic; BMI, body mass index; NAS, NAFLD Activity Score (0-8)
